# Freeze-Drying as a Novel Concentrating Method for Wastewater Detection of SARS-CoV-2

**DOI:** 10.1101/2025.01.04.25319877

**Authors:** Rui Dong, Elizabeth Noriega Landa, Hugues Ouellet, Wen-Yee Lee, Chuan Xiao

## Abstract

Detecting viral RNA from wastewater has emerged as a cost-effective approach for community-level surveillance during the recent SARS-CoV-2 pandemic. Although various concentrating methods have been developed, none are optimal for all key requirements for wastewater viral detection. Freeze-drying, a technique widely used for concentrating and preserving biological materials, remains underexplored for this purpose. This study compared the performance of freeze-drying and centrifugal ultrafiltration in terms of recovery efficiency, detection limit, and other key parameters. Early pandemic samples in this study, with extremely low viral concentrations, offered an ideal benchmark to assess their suitability for early-warning applications. Statistical analyses showed that freeze-drying achieved significantly higher recovery efficiency (0.338% ± 0.065% vs. 0.149% ± 0.046%), superior detection ratio (81.6% vs. 36.8%), and lower detection limit (0.06 vs. 0.36 copies/mL) compared to centrifugal ultrafiltration. To our knowledge, this is the first study to apply freeze-drying for wastewater-based viral detection. Despite its longer processing time, freeze-drying offers multiple advantages, including the elimination of pretreatment steps, a flexible workflow, reduced RNA degradation under cryogenic conditions, minimal pathogen exposure, lower labor demands, and less human interference during processing. These features position freeze-drying as a novel alternative for wastewater-based viral surveillance, particularly for decision-making when establishing such systems.

**Synopsis:** Freeze-drying is a new wastewater virus concentrating method that outperforms centrifugal ultrafiltration, providing a simpler, safer, and more sensitive approach for community surveillance.

## 1. Introduction

Coronavirus disease 2019 (COVID-19) is caused by the Severe Acute Respiratory Syndrome Coronavirus 2 (SARS-CoV-2), a positive-sense enveloped single-stranded RNA virus. The World Health Organization declared COVID-19 a global pandemic in March 2020. As of December 15, 2024, over 7 million deaths have been reported worldwide.^1,2^ Timely detection and monitoring of the virus at the early stages of a pandemic are crucial to mitigate its spread, and wastewater surveillance has emerged as a recognized and effective approach. Notably, in March 2020, Medema et al. reported the first detection of SARS-CoV-2 RNA in wastewater in the Netherlands, preceding major outbreaks in various regions.^3–5^ Quantifying SARS-CoV-2 RNA in sewage offers the possibility of developing an early warning system, providing valuable insights into disease prevalence and allowing public health authorities more time to intervene. Thus, sensitive and cost-effective methods are essential for such applications.

Nevertheless, assessing viral load within wastewater presents challenges due to the variable composition of sewage. Wastewater comprises both liquid and solid phases, each of which may contain viral signals.^3^ Moreover, the composition of wastewater is influenced by numerous factors, including local precipitation, drought conditions, temperature fluctuations, and water usage patterns.^4^ Therefore, the concentration of viruses in wastewater can fluctuate widely. In addition, nucleases in the environment can potentially degrade nucleic acids, particularly RNA, representing another significant factor impacting the detectable viral RNA concentration in wastewater.^5^ Considering all these variables, detecting minute quantities of specific pathogens in the complex matrix of wastewater represents a substantial challenge, necessitating a proper sample concentrating method to maximize the preservation of signal for subsequent detection.

To tackle these challenges, a concentrating method should be developed specifically to address the properties of wastewater. Given the very low viral load during early and late pandemic stages (**Figure 1a** shaded area), the recovery rate and detection limit are the most critical criteria for evaluating concentrating methods. An effective method must efficiently collect viruses from both liquid and solid phases while protecting viral genomic materials from degradation. The method should avoid (1) pretreatments such as prefiltration that could result in the loss of viruses associated with solid particles;^6,7^ (2) addition of chemicals that may inhibit downstream detection.^8–10^ Additionally, especially for RNA viruses, the approach should minimize degradation caused by ubiquitous environmental RNases.^11^ For resource-limited facilities such as wastewater treatment plants, the concentrating method should be easy to establish without specialized personnel training and biosafety infrastructure. In addition, labor-intensive processes should be minimized in the protocol for long-term surveillance applications.^12^ Despite these needs, an optimal concentrating method that effectively addresses all challenges for wastewater viral surveillance has not yet been established.

**Figure 1.**
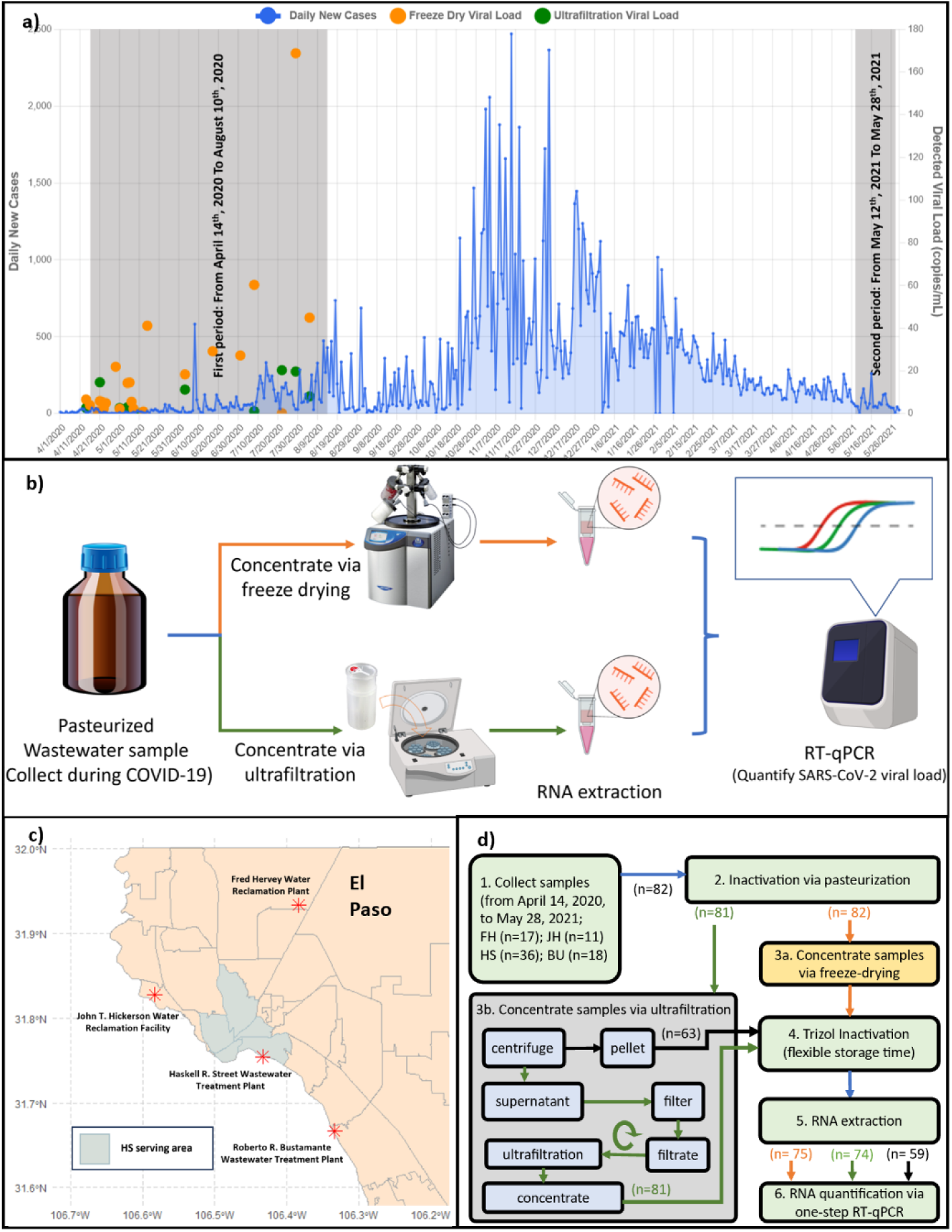
Experimental design. **(a)** El Paso COVID-19 pandemic data: daily new case numbers (blue line, left y-axis) plotted alongside detected viral loads (right y-axis, copies/mL) determined by freeze-drying (orange dots) and ultrafiltration (green dots). The two sample collection periods are shaded in grey. **(b)** Comparison workflow of concentrating methods: parallel workflow comparing freeze-drying (orange lines) versus ultrafiltration (green lines) as concentrating methods for wastewater virus detection. **(c)** Locations of wastewater treatment plants (WWTPs) in El Paso: four El Paso WWTPs marked with red “*” symbols and labeled on the map. The El Paso area is shown in pale orange, with the area served by HS shaded in grey. **(d)** Sample processing workflow and numbers: samples collected from four different WWTPs were inactivated through pasteurization and then concentrated using two methods (orange lines for freeze-drying, green lines for ultrafiltration, and black lines for pellets from the pre-clarification step of ultrafiltration) . FH, Fred Hervey Water Reclamation Plant; JH, John T. Hickerson Water Reclamation Facility; HS, Haskell R. Street Wastewater Treatment Plant; BU, Roberto R. Bustamante Wastewater Treatment Plant. The number (n) of samples processed at each step is indicated in parentheses.

The U.S. Centers for Disease Control and Prevention (CDC) has outlined five methodologies for concentrating viral particles from wastewater including (1) centrifugal ultrafiltration, (2) electronegative membrane filtration, (3) polyethylene glycol (PEG) precipitation, (4) skimmed milk flocculation, and (5) ultracentrifugation.^13^ While each of these methods employs different principles to address the challenges of low levels of viral RNA in wastewater,^6,14–19^ none fully meets all the criteria for effectively concentrating and detecting viruses. Achieving sensitive viral RNA detection in wastewater samples, especially during the onset of COVID-19 outbreak with sporadic infections, necessitates the development of improved and more robust sample concentrating methods, as existing approaches often involve trade-offs between recovery efficiency, detection sensitivity, and operational complexity.

In contrast, freeze-drying, which is not listed by the CDC, is a promising alternative concentrating method that is widely used to preserve various biological products.^20,21^ Recently, it has been applied to concentrate wastewater samples to evaluate the deactivation effect of Fe^3+^ treatment on bacteria,^22^ yet its potential for wastewater-based virus detection has not been explored. Freeze-drying effectively removes water from wastewater matrices by sublimation.

This method uniquely retains viral entities from both the liquid and solid components of wastewater, thereby enhancing detection sensitivity. Additionally, freeze-drying does not rely on reagent-dependent sample pre-treatment, thus preserving sample integrity without chemical interventions. The cryogenic environment during freeze-drying prevents the degradation of viral RNA, thereby improving the detection limit in the subsequent analyses. Although it is time-consuming to process large-volume samples, freeze-drying offers operational convenience by minimizing human oversight after initial setup. In summary, freeze-drying may represent a novel and potentially sensitive alternative method for virus detection in wastewater, as it minimizes viral loss and optimizes the protection of viral RNA. Nevertheless, its effectiveness must be assessed by comparison with existing methods.

Here we report the first application of freeze-drying to measure SARS-CoV-2 levels in wastewater. A comparative analysis was conducted between freeze-drying and the widely used centrifugal ultrafiltration as concentrating methods for wastewater samples. Throughout the subsequent sections, “ultrafiltration” will be used in lieu of the more descriptive term "centrifugal ultrafiltration" to improve conciseness. Ultrafiltration was chosen for comparison because it is among the most widely used concentrating methods for SARS-CoV-2 in wastewater and has been employed in the first^23^ and several early reports of wastewater-based surveillance^24–26^. In addition, comparative studies have demonstrated that ultrafiltration frequently achieves virus recovery efficiencies and detection sensitivities matching or exceeding those of other concentrating methods such as precipitation or flocculation, particularly at low viral loads.^12,27–29^ Given this widespread adoption and the consistently strong performance documented across multiple studies, ultrafiltration represents an appropriate benchmark against which to assess the advantages and limitations of the freeze-drying approach. Critical factors, including recovery efficiency and detection limits, were systematically compared and evaluated using RT-qPCR across both concentrating methods.

The wastewater samples tested in this study are of exceptional value because they were collected during the critical onset and waning phases of pandemic peaks (**Figure 1a**). During these periods, viral concentrations were extremely low, highlighting the value of this comparison in selecting a better concentrating method for establishing a wastewater-based early warning system. Before using these precious samples, thorough method testing and optimization were carried out using DNA and RNA standards, as well as a similar but less pathogenic coronavirus. Although each invaluable sample could be processed only once by each concentrating method due to limited sample volume, our optimized utilization strategy ensured reliable quantitative measurements while maximizing meaningful insights from these irreplaceable specimens. The resulting data provides important evidence supporting freeze-drying as a promising and innovative alternative concentrating method to detect viruses in wastewater. These findings can inform decision-making during the establishment and optimization of such monitoring systems and ultimately enhancing community-level early warning and wastewater surveillance capabilities.

## 2. Material and Methods

### 2.1. Sample Collection and Pretreatments

A total of 82 influent wastewater samples, each composited over 24 h, were collected on designated dates, from February 17, 2020, to May 28, 2021, primarily during the early stages of the first COVID-19 pandemic peak in El Paso, Texas, USA, with additional samples collected during the low and steady tail of the first outbreak (**Figure 1a** grey shaded areas, **Table S6**). The wastewater samples were collected from four wastewater treatment plants (WWTPs) in El Paso, Texas, USA. These WWTPs were Haskell Street Wastewater Plant (HS), John T. Hickerson Water Reclamation Facility (JH), Fred Hervey Water Reclamation (FH), and Roberto R. Bustamante Wastewater Plant (BU). The locations of these WWTPs are shown in **Figure 1c**.

Due to safety concerns during the early phase of the pandemic (March 2020 to July 2020), before the person-to-person transmission pathway of the SARS-CoV-2 was fully elucidated, a sample collecting protocol was developed and approved by the Institutional Biosafety Committee (IBC) of the University of Texas at El Paso (UTEP). Individuals collecting samples wore double-layer disposable nitrile gloves, splash goggles, N95 or surgical masks, and disposable lab coats as a precautionary measure. The designs of our wastewater sample collection and processing protocols in the following sections reflect a strong commitment to high safety standards in adhering to the stringent biosafety standards set by institutions such as the CDC and NIH for emerging unknown pathogens. This proactive approach to biosafety was crucial during uncertain times. Nevertheless, these precautionary sample collection protocols may also be applicable for future emerging viruses.

Due to limited storage space in −80 °C freezers, a maximum of approximately 500 mL of wastewater could be collected for each sampling event, and each sample was split into two ∼200 mL portions, which were analyzed in parallel by the two concentrating methods: freeze-drying and ultrafiltration (**Figure 1b**). During the early period from February to August 2020, 68 samples were collected and stored at −80 °C using the IBC-approved biosafety protocol developed at UTEP, prior to finalization and IBC approval of the analytical protocol at the end of 2020. Some samples from certain WWTPs were unavailable due to technical and personnel challenges encountered during the pandemic.

Due to the limited lab accessibility during the pandemic, human resource constraints, and the restricted storage space in −80 °C freezers, sample collection was paused between August 2020 and April 2021. After samples were processed using the fully IBC-approved protocol, freeing storage space in −80 °C freezers, sample collection resumed in May 2021. The collection frequency during the second period was more sporadic, depending on the processing speed of samples stored. For the second period, a total of 14 samples were collected.

Samples were collected in pre-cleaned, wide-mouth, sterile leak-proof bottles (Fisherbrand^TM^, Cat# 05719709), transported in double Ziploc bags within hard-plastic coolers to the biosafety level 2 (BSL-2) lab, and handled under standard biosafety and disinfection procedures. For early pandemic samples, from April 2020 to September 2020, bottles were immediately frozen and stored at −80 °C. Later, after CDC published its guideline in September 2020 to inactivate pathogens for wastewater virus detection,^30^ samples collected after September 2020 were pasteurized in a water bath at 65 °C for 90 min before being stored at −80 °C. For those early samples without pasteurization, before processing, they were thawed and pasteurized as recommended by CDC guidelines. Following pasteurization, 82 samples were concentrated using freeze-drying and 81 using ultrafiltration, respectively.

### 2.2. Wastewater Sample Concentrating via Freeze-drying

Although samples were pasteurized and thus considered to be BSL-1 according to the CDC guidelines,^30^ a fully sealed freeze-drying setup was cautiously designed and implemented using a FreeZone 2.5 Liter −84 °C Benchtop Freeze Dryer (Labconco™, Cat# 710201000). This design is illustrated in **Figure 2**. The biosafety setup incorporated multiple safeguards: (1) The freeze dryer vacuum chamber contained PTFE-coated coil, which is resistant to erosive disinfectants used after each freeze-drying cycle; (2) Three layers of Filter papers (Labconco™, Cat# 7544810) and an aseptic valve (Labconco™, Cat# 7543900) were added between the fast-freeze flask and manifold to prevent any potential accidental pathogen from passing through the system; (3) A HEPA filter was added before the vacuum pump as the last filter to capture any potential accidental pathogen leakage and spillage; (4) Wastewater samples were placed inside plastic containers which were wrapped with absorbent pads in Fast-Freeze Flasks. The absorbent pads were intended to capture any accidental spills; and (5) The exhaust pipe of the vacuum pump was connected to a chemical fume hood, to exhaust odorous volatile organic compounds from the wastewater. Even with these safety designs, as an additional precaution, all potentially contaminated containers and the flow path before the HEPA filter were disinfected after each freeze-drying run.

**Figure 2.**
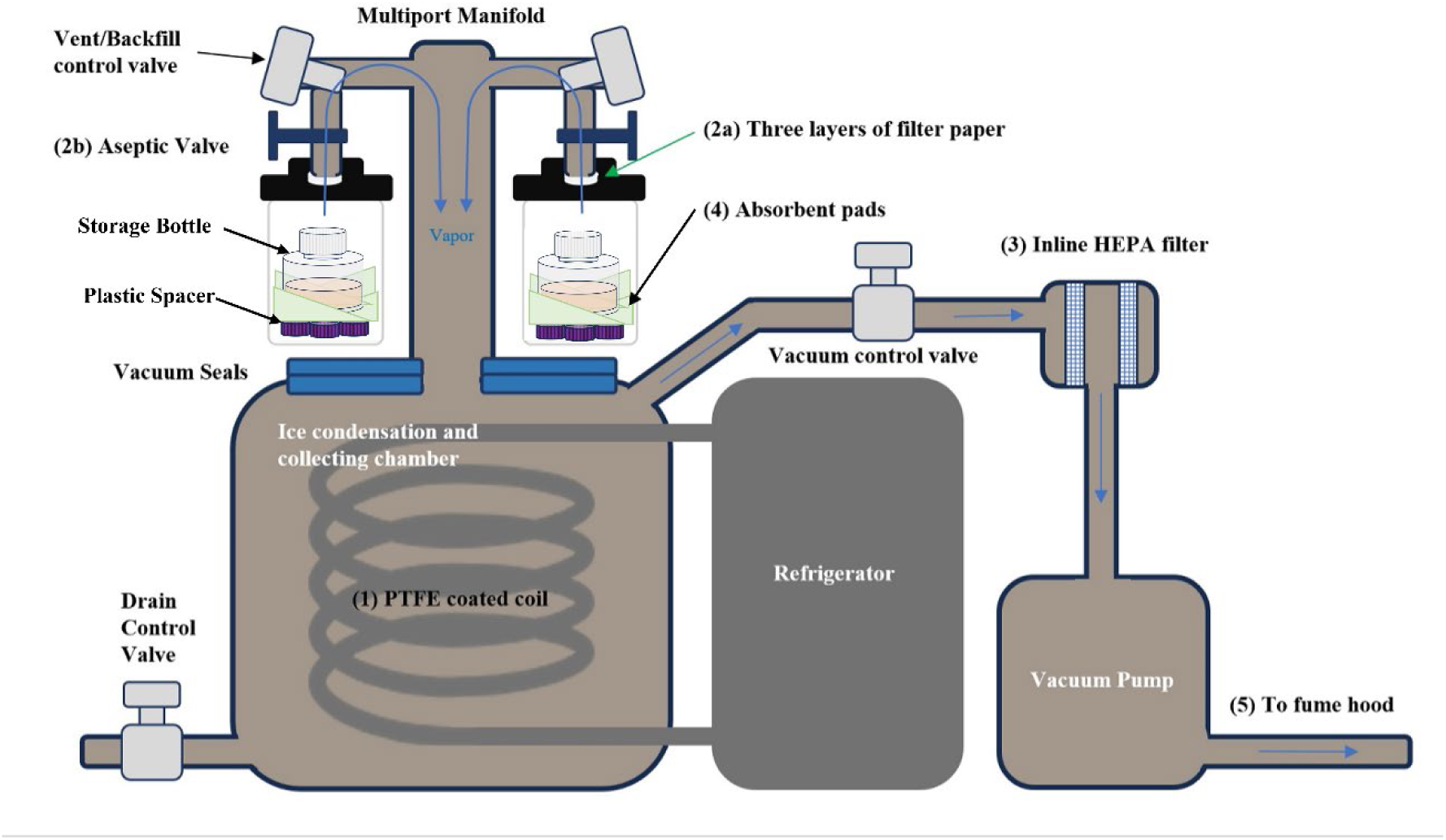
Freeze-drying setup with biosafety safeguards. Key biosafety safeguard features are highlighted and labeled with numbers in the diagram.

Systematically varied parameters such as sample volume, sample container (flask or bottle), wrapping or no wrapping, distance between the sample container and freeze-drying flask, the type of freeze-drying flask, and freeze-drying time were tested over several months before optimized freeze-drying conditions were established as described below.

As shown in **Figure 1b and d**, prior to the freeze-drying procedure, approximately 200 mL of each pasteurized wastewater sample from **Section 2.1** was weighed and aliquoted into a 250 mL sterile plastic storage bottle (Thermo Scientific, Cat# 4550250), wrapped with absorbent pads, placed into a Ziploc bag, and frozen at −80 °C at least one night.

On the day of concentrating, the frozen samples in storage bottles wrapped with absorbent pads were then placed in 600 mL Complete Fast-Freeze Flasks (Labconco™, Cat# 7540800) with storage bottle caps loosened on top of plastic spacer (three caps from 50 mL conical tubes) (**Figure 2**). Freeze-drying was performed at −86 °C and 0.040 mbar. After approximately three days, sample masses were reduced to about 3 g, with a small amount of ice remaining visible.

Complete desiccation was not pursued, as it would substantially increase processing time and make the dried material more difficult to resolubilize in the subsequent step. Volume reduction factors were estimated from sample weights measured before and after freeze-drying.

The freeze-dried samples were dissolved in three volumes (v/w; ∼9 mL) of TRIzol™ LS reagent (Thermo Fisher Scientific, Cat# 10296-028). Samples were vortexed to ensure thorough mixing before storing at −20 °C until viral RNA extraction. It has been proven that TRIzol deactivates all viral pathogens such as SARS-CoV-2 in the sample.^31^ Therefore, the following steps were considered as BSL-1.

### 2.3. Sample Concentrating via Ultrafiltration

In parallel with concentrating samples via freeze-drying, another ∼200 mL of pasteurized wastewater sample from **Section 2.1** was aliquoted into four 50 mL conical tubes and loaded into swinging buckets (Thermo Fisher Scientific, Cat# 75003655) with adaptors (Thermo Fisher Scientific, Cat# 75003683) for the TX-400 rotor (Thermo Fisher Scientific, Cat# 75003181). The swinging buckets were balanced and sealed with ClickSeal Biocontainment Lids (Thermo Fisher Scientific, Cat# 75003656) and wiped with disinfectant on the outside before centrifuging at 3,000 × *g* for 15 min using Multifuge X1R Refrigerated Centrifuge (Thermo Fisher Scientific, Cat# 75004251). After centrifugation, the supernatant was filtered using a 0.22 µm PES bottle filter (Corning™, Cat# 431096) while the pellets were collected and weighed. In total, 63 pellets were collected from 70 ultrafiltration samples (**Figure 1d**). Three volumes of TRIzol™ LS reagent relative to the measured pellet weight were added to the pellet. TRIzol treated pellets were vortexed to ensure thorough mixing before being stored at −20 °C until viral RNA extraction to assess whether viral signals were associated with the solid phase. All these procedures were carried out in a biosafety cabinet (BSC) to ensure safety. **Figure S1** illustrates the filter system with multiple, precautionary safeguards: (1) A trap bottle with disinfectant at the bottom was connected to the filter bottle to capture any potential accidental pathogen leakage and spillage; (2) An in-line 0.22 µm filter was added before the vacuum valve to capture any potential accidental pathogen leakage and spillage.

Inside the BSC, the filtered supernatants from the previous step were aliquoted (approximately 70 mL) into ultrafiltration units. In this study, Centricon Plus-70 with 10 kDa MW cutoff membrane (MilliporeSigma, Cat# UFC701008 for 10 kDa) were used for most wastewater samples. Two Centricon Plus-70 units with a 100 kDa cutoff membrane (MilliporeSigma, Cat# UFC71008 for 100 kDa) and four 15 mL Amicon Ultra Centrifugal Filter with a 10 kDa cutoff membrane (MilliporeSigma, Cat# UFC901024) were used in spiked recovery experiments to evaluate the impact of different filter types. These concentrators were then placed into disinfected swinging buckets (Thermo Fisher Scientific, Cat# 75003655, with seal cap Cat# 75003656). The buckets were carefully balanced and completely sealed with ClickSeal Biocontainment Lids (Thermo Fisher Scientific, Cat# 75003656) and wiped with disinfectant on the outside before being taken out of the BSC for loading onto the TX-400 rotor (Thermo Fisher Scientific, Cat# 75003181).

The samples were centrifuged at 3,500 × *g* for approximately 15 min using Multifuge X1R Refrigerated Centrifuge (Thermo Fisher Scientific, Cat# 75004251). After centrifugation, the swinging buckets were taken back into the BSC, and the passthrough liquid was disinfected and discarded. Following the same centrifugation protocol, the remaining filtered supernatants were added to Centricon units and carefully balanced for the next round of concentrating via ultrafiltration.

After three to four rounds of centrifugation, depending on wastewater viscosity and matrix composition (e.g., dissolved high-molecular-weight organics and residual colloids), the sample volume was reduced from 200 mL to approximately 0.5 mL. Following the manufacturer’s instructions, the Centricon Plus-70 filters were inverted and centrifuged at 1,000 × *g* to collect the concentrates. The concentrated samples were subsequently transferred to new tubes and mixed with three volumes (∼1.5 mL) of TRIzol™ LS reagent. TRIzol treated samples were vortexed to ensure thorough mixing before being stored at −20 °C until viral RNA extraction. Volume reduction factors for ultrafiltration were determined by comparing the weights of the samples before and after concentrating. All the filters and liquid transfer pipettes were soaked in ∼1% bleach for at least 15 min before being safely discarded.

### 2.4. Primer–Probe Selection and Standard Curve Development

As suggested by the CDC, SARS-CoV-2 RNA was quantified by one-step RT–qPCR using N1 and N2 primer-probe sets (**Table S1**) purchased from IDT (Cat# 10006713).^32^ The performance and efficiency of N1 and N2 primer-probe sets in the RT-qPCR assay were first evaluated by generating standard curves using 10-fold serial dilutions (1 to 10^5^ copies per reaction) of templates: (1) a DNA plasmid containing the complete nucleocapsid gene of SARS-CoV-2 (IDT, Cat# 10006625), and (2) Twist synthetic SARS-CoV-2 RNA (Twist Bioscience, Cat# MT0075s44) with stock concentration of 2.0*×*10^5^ copies/μL and 1.0*×*10^6^ copies/μL, respectively.

Each diluted standard was aliquoted in triplicate and analyzed using the TaqPath™ 1-Step RT-qPCR Master Mix (Thermo Fisher Scientific, Cat# A15299) in 20 µL reactions containing 5 µL template, with primer and probe concentrations listed in **Table S1**. Triplicate water samples were included as the negative controls. The one step RT-qPCR reactions were run at 50 °C for 15 min and 95 °C for 2 min, followed by 45 cycles of 95 °C for 5 s and 55 °C for 30 s, per the manufacturer’s recommendations.

Standard curves were generated by least-squares regression of technical replicate data points, and the fitting quality (R²) and assay efficiency (E) were evaluated (**Figure S2; Table S2**). To test template stability, standard curves were also generated after storing of standards at −20 °C for up to four months. After evaluation (**Section S3; Section 3.1**), the N1 primer-probe set with the DNA standard was selected to quantify all extracted RNA from wastewater samples.

### 2.5. Viral RNA Extraction and Quantification

SARS-CoV-2 RNA was extracted from the concentrated samples and purified using the PureLink^TM^ RNA Mini Kit (Thermo Fisher Scientific, Cat# 12183018A). Prior to RNA extraction, the frozen samples mixed with TRIzol were thawed at room temperature. For every milliliter of sample mixed with TRIzol, 0.2 mL Chloroform was added. The mixture was then centrifuged at 12,000 × *g* at 4 °C for 15 min using a Microfuge 20R (Beckman Coulter, Cat# B30148). Following centrifugation, the colorless upper aqueous phase, which contains the RNA, was carefully extracted according to the manufacturer’s instructions. The bound RNA was eluted from the spin cartridge in 30 µL of ribonuclease-free water and collected by centrifugation. To maximize RNA recovery, a second elution was performed by adding 100 μL of ribonuclease-free water and centrifuging. The purified viral RNA was either immediately analyzed by one-step RT-qPCR or stored at −20 °C to minimize degradation, depending on thermocycler availability.

RT-qPCR assays for RNA extracted from the same sampling event but split and concentrated in parallel using the two methods, were always performed in the same runs to ensure identical post-extraction processing.

Each RNA elution from extraction was aliquoted in triplicate and analyzed under identical one-step RT-qPCR conditions as the DNA standard using the N1 primer-probe set (**Section 2.4**).

Viral concentrations were calculated as described in **Section S4**. These values represent the virus recovered by each concentrating method and therefore allow direct comparison of their analytical sensitivity.

During method development, including primer-probe set and standard evaluations, a Magnetic Induction Cycler (Bio Molecular Systems, Cat# 71-101; 48 individual-tube reactions) was used. After optimization and standardization, a StepOnePlus™ Real-Time PCR System (Thermo Fisher Scientific, Cat# 4376600; 96-well plate format) was employed using identical thermocycling conditions for all of the wastewater samples. DNA standards were run on both thermocyclers (**Figure S2c**), and sample quantification was based on the standard curve generated on the same instrument. Unfortunately, RT-qPCR data for some samples were lost due to an unexpected crash of the computer controlling the StepOnePlus™ Real-Time PCR System. Ultimately, quantification data were obtained from 75 freeze-dried samples, 74 ultrafiltration samples, and 59 pellets collected during the pre-clarification step of ultrafiltration (**Figure 1d; Table S6**).

The one-step RT-qPCR assay was programmed for 45 amplification cycles with Ct values up to 42 considered positive based on consistent amplification in technical replicates. This practice aligns with CDC peer-reviewed SARS-CoV-2 wastewater surveillance protocols using 45-cycle RT-qPCR, which routinely accept late Ct values near the cycling limit as positive when supported by replicate consistency^33^ and used by literature.^34,35^

### 2.6. Recovery Efficiency Determination Using Human Coronavirus OC43

Human coronavirus OC43 was used as a surrogate virus to compare recovery efficiencies of the two concentrating methods. Both OC43 and SARS-CoV-2 are beta-coronaviruses that infect the human respiratory tract^36,37^, but OC43 causes only mild disease and therefore can be handled in laboratory recovery experiments with fewer biosafety concerns.^38,39^

A standard curve of heat-inactivated OC43 (ZeptoMetrix, Cat# 0810024CFHI) was generated to estimate the amount of virus spiked into wastewater. The stock concentration was provided in TCID_50_/mL, an infectivity-based unit defined as the dilution required to infect 50% of cell monolayers.^40^ Although TCID_50_ does not directly represent genome copy number, it remains proportional to infectious virus abundance. Because the objective of this study was method comparison rather than absolute RNA quantification, TCID_50_ provided a consistent internal reference and has been used in previous wastewater virus studies.^41–44^ All standard curve preparation and spiking experiments used the same OC43 stock, and recovery efficiency was expressed as a unitless ratio, making TCID_50_ appropriate for this framework.

An exact 100 μL aliquot of inactivated OC43 (5.0 × 10⁵ TCID_50_/mL) was serially diluted tenfold using molecular-grade water. Dilutions were subjected to the same TRIzol treatment, RNA extraction, and RT-qPCR procedures described previously, using an OC43-specific primer-probe set^45^ (**Table S1**). A standard curve was established by plotting Ct values against the logarithm of OC43 concentration (TCID_50_/mL). Although extraction efficiency from stock dilutions is unlikely to be 100%, both spiked wastewater samples and OC43 standards underwent identical extraction and purification procedures, allowing proportional comparison.

For recovery efficiency experiments, 5.0 × 10³ (low) or 2.5 × 10⁴ (high) TCID_50_ of heat-inactivated OC43 were spiked into 200 mL raw influent wastewater (10 μL or 50 μL of the stock solution, respectively). A 1 mL aliquot of the spiked sample was immediately mixed with 3 mL TRIzol LS reagent (F-1mL and U-1mL in **Table S3**). The remaining sample, along with two additional unspiked 200 mL controls (F0 and U0), was concentrated using either freeze-drying or ultrafiltration following the procedures described in **Sections 2.2** and **2.3**, and viral RNA was quantified by RT-qPCR in technical triplicate (**Section 2.5**).

For low-dose experiments (**Figure 3**; **Table S3**), one sample for each concentrating method was spiked after pasteurization (F4 and U4) to evaluate potential pasteurization effects. Three freeze-drying samples (F5–F7) and four ultrafiltration samples (U5–U8) were spiked before pasteurization. To examine the influence of filter type under low-dose conditions, ultrafiltration samples used different filters: Centricon Plus-70 with 10 kDa cutoff (U5), Centricon Plus-70 with 100 kDa cutoff (U6 and U7), and four Amicon Ultra 15 mL centrifugal filters with 10 kDa cutoff (U8).

**Figure 3.**
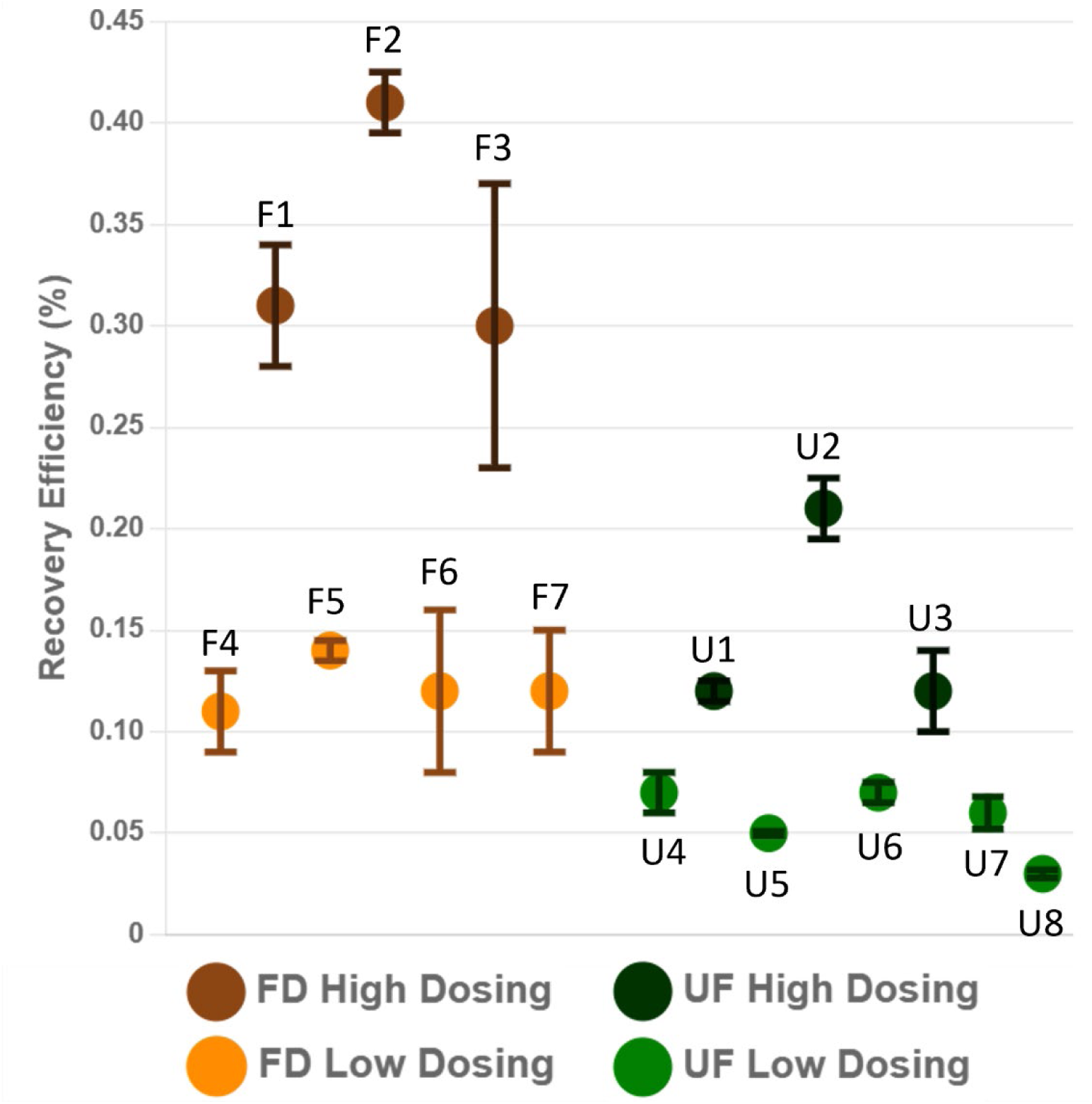
Recovery efficiency of the OC43 surrogate virus using freeze-drying and ultrafiltration concentrating methods. Heat-inactivated OC43 was spiked into influent wastewater samples, and recovery efficiencies were calculated based on RT-qPCR quantification of recovered viral RNA relative to the amount initially spiked. Sample IDs correspond to individual experimental replicates. Orange markers represent freeze-drying samples, and green markers represent ultrafiltration samples. Darker colors (F1–F3 and U1–U3) indicate higher spike levels, whereas the lighter-colored markers indicate lower spike levels. For selected samples, OC43 was added before pasteurization for most samples, except F4 and U4, which were spiked after pasteurization. For ultrafiltration samples, U1–U5 used 70 mL Centricon Plus-70 units with a 10 kDa cutoff membrane; U6 and U7 used 70 mL Centricon Plus-70 units with a 100 kDa cutoff membrane; and U8 used four 15 mL Amicon Ultra centrifugal filter units with a 10 kDa cutoff membrane. Error bars represent variability among technical RT-qPCR replicates.

For high-dose experiments, three independent wastewater samples (biological n = 3) were spiked for each concentrating method (F1–F3 and U1–U3 in **Figure 3** and **Table S3**).

Ultrafiltration in these experiments used Centricon Plus-70 with a 10 kDa cutoff membrane. Recovered virus concentrations were estimated from RT-qPCR Ct values and the OC43 standard curve, expressed as TCID_50_/mL, and used to calculate total recovered virus based on sample volumes. Recovery efficiency was defined as the ratio of recovered virus to the amount initially spiked (**Figure 3**; **Table S3**; **Table S4** for statistical analyses).

Recovery efficiencies were determined from at least three independent biological replicates (n ≥ 3) per method, each analyzed in technical triplicate, allowing calculation of the mean and standard deviation. Molecular-grade water blanks confirmed the absence of contamination. A Wilcoxon rank-sum test, which does not assume normal distribution,^46–48^ was used to evaluate statistical differences between the two concentrating methods. The statistical analyses were conducted with functions wilcox.test() from the stats (version 4.3.1) package in R Studio.^49^

### 2.7. Method-specific Detection Limits

Method-specific detection limits were operationally defined as the lowest non-zero SARS-CoV-2 concentration (copies/mL) detected among processed wastewater samples, reflecting practical sensitivity in complex matrices. This approach has been used in previous wastewater studies.^50,51^ Method-specific detection limits are reported in the last row of **Table S6**. Additional spiking experiments using DNA standards at the estimated threshold were performed to partially verify the magnitude level of these limits (**Section S7; Table S5**). These values represent operational metrics for comparing workflow sensitivity rather than formal Minimum Information for Publication of Quantitative Real-Time PCR Experiments (MIQE)-compliant assay limits of detection (LODs).^52,53^

### 2.8. COVID-19 Case Data and Statistical Analyses

Daily COVID-19 records from April 14, 2020, to May 28, 2021, in El Paso, TX, were collected from https://www.epstrong.org/results.php and https://elpasocovid19tracker.com/index.html (accessed 2025-11-18) (**Figure 1a**). Viral load data from Haskell Street Wastewater Plant (HS), which had the largest number of wastewater samples, together with the corresponding daily COVID-19 case counts on the wastewater sample collection dates were compiled and listed in **Table S7**. The areas from where the case numbers were collected correspond to the regions served by HS, including zip codes 79901, 79902, 79903, 79905, and 79930 (**Figure 1c**).

The association between total COVID-19 cases and SARS-CoV-2 viral load in wastewater was first tested for normality using the Shapiro–Wilk test.^54^ The variables significantly deviated from normality (p < 0.05). Because Pearson’s correlation assumes normally distributed data, Spearman’s rank correlation (ρ), a nonparametric test that does not require normality, was instead used to assess the relationship between the variables.^55^ This correlation can be denoted as rho “ρ”, ranging from -1 to 1, provided a quantitative measure of the observed relationship between the two variables: daily COVID-19 case numbers and SARS-CoV-2 viral load in wastewater. A ρ close to zero indicates no correlation, while close to 1 and -1 indicates strong positive or negative correlation, respectively. The data processing and statistical analyses were conducted with functions shapiro.test(), cor.test(), and wilcox.test() from the stats (version 4.3.1) package in R Studio.^49^

## 3. Results and Discussion

### 3.1. RT-qPCR Primer-Probe Set Selection and Template Stabilities

Based on the evaluation of primer–probe sets and DNA/RNA standards, the N1 primer-probe set was selected as the optimal choice for RT-qPCR quantification of wastewater samples due to its stronger linear correlation and superior amplification efficiency, whereas DNA standards were chosen because of their greater stability (**Section S3**). While RNA standards are preferred for absolute quantification of RNA viruses due to minor differences in RT-qPCR amplification efficiency between DNA and RNA templates, synthetic RNA standards are costly, prone to degradation, and less stable for long-term usage. DNA standards have been widely used in SARS-CoV-2 wastewater studies^56,57^ and were empirically validated in this study, which focusing on comparing to different concentrating methods. Our primary conclusions regarding relative method performance (recovery efficiency, detection ratio, and method-specific detection limit) are unaffected by the choice of standard, as they are based on method-paired analyses in which identical standards were applied to both freeze-drying and ultrafiltration workflows.

Based on the observed degradation of RNA standards (**Section S3, Figure S2**), specific precautions must be followed to mitigate RNA degradation in concentrated wastewater and preserve its integrity for potential future analyses. Immediate sample processing after collection is ideal for maximizing RNA extraction yield and maintaining its integrity. However, logistical constraints often necessitate sample storage, for which TRIzol^TM^ or similar denaturing reagents are highly recommended. TRIzol effectively inhibits RNase activity, thereby preventing RNA degradation.^58^ Concentrated samples in TRIzol demonstrate superior stability for long-term storage, particularly when stored at −80 °C.^59^ Thus, maintaining samples in TRIzol without RNA extraction until imminent RT-qPCR analysis is considered the optimal storage strategy.

Moreover, minimizing freeze-thaw cycles of samples, regardless of the storage method, is critical to prevent RNA degradation. RNases are prevalent and can regain enzymatic activity once denaturing conditions are removed.^60^ Consequently, after RNA is extracted from the TRIzol storage solution, downstream procedures should be performed promptly to minimize RNA degradation and ensure the reliability and accuracy of the subsequent RT-qPCR analyses.

It is important to acknowledge that logistical constraints during the study period led to some variation in sample processing. Specifically, some samples were stored at −80 °C prior to pasteurization, others were pasteurized and then stored, and some underwent immediate pasteurization followed by extraction (**Section 2.1**). However, for each methodological comparison between freeze-drying and ultrafiltration, paired samples were subjected to identical pre- and post-concentrating steps. This internal consistency ensures that the observed differences are attributable to the concentrating methods themselves.

### 3.2. Comparison of Recovery Efficiency Between Two Concentrating Methods

Recovery efficiencies (**Section 2.6** and **S5**) were 0.338% ± 0.065% for freeze-drying and 0.149% ± 0.046% for ultrafiltration at a high spike level (2.5 × 10^4^ TCID_50_ per 200 mL). At a lower spike level (5 × 10^3^ TCID_50_ per 200 mL), efficiencies decreased to 0.123% ± 0.025% and 0.062% ± 0.011%, respectively. Freeze-drying showed significantly higher recovery than ultrafiltration (Wilcoxon rank-sum test, p = 5.7 × 10⁻⁴ for the high dose and p = 1.7 × 10^⁻5^ for the low dose) **(Table S4**).

The recovery efficiency observed in this study is lower than the ∼25% reported by Ye et al. using ultrafiltration,^29^ likely due to the substantially higher spiking concentration in that study (∼3 × 10⁴ PFU/mL). In contrast, our highest spiking condition corresponded to 125 TCID_50_/mL, equivalent to approximately 47–58 PFU/mL based on established theoretical61 and simulation62 relationships (0.56–0.69 PFU per TCID_50_), representing a >500-fold lower input concentration.

Lower input concentrations are expected to yield reduced apparent recovery efficiencies due to stochastic effects and increased influence of matrix-associated losses. Consistent with this expectation, our two-level spiking experiments showed higher recovery efficiencies at higher initial concentrations (**Figure S3; Table S4**).

Based on literature-reported relationships between genome copy number and TCID_50_ for the OC43 stock,^61^ together with our measured recovery efficiency and RT-qPCR standard curves (Figure S2; Table S3), the expected Ct value is approximately ∼28, which is in reasonable agreement with the observed values (**Table S3**). However, the ratio between infectivity-based units and genome copy number is inherently variable and depends on virus preparation, inactivation, and environmental conditions. Therefore, this conversion is used only as an approximate scaling factor for consistency checks rather than as an absolute equivalence.

RNA extracted from independently spiked 1 mL samples without concentration (F-1mL and U-1mL in **Table S3**) and from concentrated 200 mL unspiked wastewater samFples (F0 and U0 in **Table S3**) showed no detectable Ct values within 45 cycles, indicating negligible endogenous OC43 signals. Recovery efficiencies were not significantly affected by spiking before or after pasteurization (p = 0.27 for freeze-drying; p = 0.11 for ultrafiltration, **Table S4**).

For ultrafiltration, four 15 mL Amicon Ultra filters yielded lower recovery than a single 70 mL Centricon Plus-70 unit (p = 0.004), whereas membrane cutoff (10 kDa vs 100 kDa) had no significant effect within 70 mL filters (p = 0.47) (**Table S4**). Accordingly, data from 15 mL filters were excluded from ultrafiltration averages.

### 3.3. Comparison of Early Detection of Two Concentrating Methods

The materials used in this study were precious and required careful stewardship. Prior to processing the pandemic-period samples, multiple trials using non-pandemic wastewater were conducted to optimize the workflow and quantification procedures. Due to the limited volumes collected during the early pandemic period, each invaluable sample could only be processed only once per concentrating method. However, triplicate RT-qPCR analyses were performed on RNA extracted from the concentrated samples, thereby maximizing the use of each collection and enabling reliable quantitative measurements despite constraints in sample availability, logistical challenges, and restricted laboratory access during pandemic-related shutdowns.

To evaluate the detection sensitivity of the freeze-drying and ultrafiltration methods, wastewater viral load data from the early onset phase of the COVID-19 pandemic (April to May 2020) were analyzed. A total of 38 samples were collected from wastewater treatment plants in El Paso, Texas, USA (**Table S6**, shaded rows), of which 33 had RT-qPCR data available. Using only the first elution, freeze-drying identified 31 positive samples (93.9%), whereas ultrafiltration detected 14 (42.4%). When the second elution was included, the detection rate for freeze-drying increased to 32 positives (97.0%), while ultrafiltration showed no increase (**Table 1**). The significantly higher detection rate observed with freeze-drying (p = 1.01 × 10⁻⁵ using only the first elution; p = 1.36 × 10⁻⁶ when second elutions were included, **Section S6**) highlights its superior ability to concentrate viral material compared with ultrafiltration. This advantage is particularly important for early warning systems for emerging virus surveillance.

**Table 1.** Positive detection rate and viral load for wastewater samples collected from four WWTPs between April and May 2020.

| Site | Number of Sample | Number of Positive Sample |  |  |  | Average Viral Load (copies/mL) |  |
| --- | --- | --- | --- | --- | --- | --- | --- |
|  |  | Freeze-drying | Ultrafiltration | Freeze-drying | Ultrafiltration | Freeze-drying | Ultrafiltration |
| HS | 16 | 14 (15)* | 5 | 87.5 (93.8)* | 31.3 | 8.35 | 1.45 |
| JH | 1 | 1 | 1 | 100.0 | 100.0 | 6.76 | 3.08 |
| FH | 6 | 6 | 5 | 100.0 | 83.5 | 7.54 | 1.68 |
| BU | 10 | 10 | 3 | 100.0 | 30.0 | 12.43 | 2.31 |
| Total | 33 | 31 (32)* | 14 | 93.9 (97.0)* | 42.4 | $p=1.01 \times 10^{-5}$ ($1.36 \times 10^{-6}$) | |

\* Numbers in the parentheses are values obtained from additional positive samples when second elution was included.

### 3.4. Comparison of Viral Concentration Detected via the Two Concentrating Methods

The superior detection sensitivity of the freeze-drying method was further supported by the higher virus load obtained during RT-qPCR analyses. Across all four wastewater treatment plants, the freeze-drying method consistently exhibited a higher amount of viral RNA compared to ultrafiltration (**Table S6**). A Wilcoxon rank-sum test, a nonparametric test that compares two independent groups without assumption of normal distribution,^46^ was conducted to compare virus loads in the original wastewater samples across all four facilities. As described in **Section 2.5** and **S4**, the total virus copies were estimated using the DNA standard curve and the Ct values. For both freeze-dry and ultrafiltration methods, both first and second elutions were combined to calculate the total RNA copies, which can then be calculated to the detected viral load (copies/mL) in the original wastewater samples. Where a method or elution did not detect the virus, it was recorded as zero copies. Across 74 paired samples, freeze-drying detected a significantly higher median viral load (7.05 copies/mL [IQR 3.52-31.17]) than ultrafiltration (1.19 copies/mL [IQR 0.00-5.49]) (**Table S6,** bottom rows) (Wilcoxon rank-sum test, p = 2.39 × 10⁻⁹).

### 3.5. Comparison of Method-Specific Detection Limits of the Two Concentrating Methods

Using the definition in **Section 2.7**, the freeze-drying method exhibited a lower method-specific detection limit (0.06 copies/mL) than ultrafiltration (0.36 copies/mL) (**Table S6**, last row). The corresponding Ct values (39.2 and 36.6) fall within the validated range of the standard curve, supporting the validity of these measurements.

DNA standard spiking experiments yielded a detectable range of 40–80 copies per 200 mL (**Section S7**, **Table S5**). Although this appears higher than the 12 copies per 200 mL (0.06 copies/mL) inferred from wastewater samples, this difference is expected given the low recovery efficiencies observed in this study (0.4%–0.1%; **Table S3**). Accounting for recovery losses, detection of 12 copies after concentration corresponds to approximately 3,000–12,000 copies in the original sample, indicating that the apparent discrepancy arises from recovery constraints rather than overestimation of sensitivity.

Such discrepancies are expected, as wastewater detection sensitivity is strongly influenced by method-dependent recovery efficiency and can vary by several orders of magnitude across workflows.^62^ Importantly, the spiking experiments do not constitute a full MIQE-compliant determination of assay LOD; rather, they provide partial verification of the order of magnitude of the method-specific detection limits defined here.

### 3.6. Matrix Associated Inhibitory Factors

Among the 75 wastewater samples analyzed, 73 freeze-dried samples were positive in either the first or second elution. Of these, 68 (93.1%) showed detectable viral signals in the second elution, including 25 (34.3%) samples with lower Ct values than in the first elution; notably, 7 of these were undetected in the first elution (**Table S6**). Similarly but slightly less, among the 47 positive samples out of 74 ultrafiltration samples, only 33 (70.2%) showed detectable viral signals in the second elution, and 10 (21.3%) exhibited lower Ct values in the second elution compared with the first; 5 of these were undetected in the first elution.

This phenomenon may occur because both concentrating methods can potentially co-concentrate RT-qPCR inhibitors. Many of these inhibitory substances may be enriched in the first elution, while the remaining inhibitors become further diluted in the second elution (100 µL in the second elution vs 30 µL in the first), leading to similar or even lower Ct values. Because the freeze-drying process concentrates all solutes, including those associated with the solid phase, inhibitory substances that affect RT-qPCR may also be enriched to a greater extent than in ultrafiltration.

Although freeze-drying showed a higher percentage of detectable viral signals in the second elution, which could lead to greater underestimation of viral abundance compared with ultrafiltration, our data consistently showed lower Ct values and higher detection ratios in freeze-dried samples than in ultrafiltration samples. This indicates that any inhibitory effects were insufficient to offset the gain in viral recovery.

Unfortunately, this potential inhibition phenomenon was recognized only after completion of all RT-qPCR runs, and retrospective inhibition testing was not feasible due to limited remaining sample volume. Nevertheless, systematic inhibition assessments and the incorporation of inhibitor-removal steps^63^ should be explored in future optimization of freeze-drying as a wastewater virus concentration method.

### 3.7. Correlation Between Wastewater Viral Load and COVID-19 Case Numbers

The relationship between wastewater viral load and community COVID-19 cases was examined to assess the epidemiological relevance of the two concentrating methods. Viral loads calculated from samples processed by freeze-drying and ultrafiltration (**Table S7**) were analyzed separately against reported COVID-19 case numbers in El Paso, TX, USA.

Normalization of viral load to account for variations in human wastewater contribution is often recommended. Creatinine-based normalization was attempted using a commercial colorimetric assay (**Section S8**);^64,65^ however, creatinine concentrations in the collected samples were consistently below the detection limit of the kit (**Table S6,** last column), preventing reliable normalization as seen in literature.^64^ Therefore, subsequent analyses were conducted using unnormalized viral load data, and the results are presented only as supplemental observations rather than definitive epidemiological conclusions.

Shapiro–Wilk tests indicated that viral load and case data were not normally distributed (p = 4. 8 × 10^-7^, 1.8 × 10-6, and 1.4 × 10^-7^, for virus concentration detected by freeze-drying, those detected by ultrafiltration, and community COVID-19 case numbers, respectively),^54^ and therefore Spearman rank correlation^55^ was used to evaluate associations between wastewater viral load and daily reported COVID-19 cases. Active case numbers were unavailable during the early pandemic period (April–May 2020), so daily reported cases were used instead.

Correlation analysis showed a weak positive but non-significant relationship between viral load measured by freeze-drying and daily cases (ρ = 0.209, p = 0.339). Similarly, viral loads obtained by ultrafiltration showed a moderate but non-significant correlation with reported cases (ρ = 0.379, p = 0.0746). These results are broadly consistent with the variable correlations reported in previous wastewater surveillance studies.^23,66,67^ However, interpretation is limited by the small sample size and the lack of reliable normalization for human wastewater contribution and other factors.^68^

### 3.8. Initial Capital Investment and Operational Costs

From the perspective of initial capital investment, a freeze-dryer is comparably priced to the refrigerated centrifuge required for ultrafiltration. The freeze-dryer used in this study costs approximately $14,000, whereas the centrifuge costs approximately $16,000. Freeze-drying additionally requires an ultra-low temperature (ULT) freezer, even lower-cost undercounter models (e.g., K2 Scientific, Cat. #K203ULT) are approximately $7,200. However, most laboratories already maintain ULT freezers for biological sample preservation, reducing the need for this additional investment. In terms of operational consumables, ultrafiltration requires disposable filters such as the filter used mostly in this study (MilliporeSigma Cat. #UFC701008) cost approximately $43 per sample, whereas the freeze-drying method uses only inexpensive sample bottles (Thermo Scientific Cat. #4550250) costing approximately $2.4 per sample. All equipment and consumable prices list above were obtained with an institutional educational discount at the time of the study.

### 3.9. Pretreatment

Prior to concentrating, wastewater samples were pasteurized, a standard biosafety practice in SARS-CoV-2 wastewater analysis to inactivate infectious virus. Some studies have reported that pasteurization may enhance SARS-CoV-2 RNA detection by disrupting particulate matrices and releasing particle-associated viral RNA.^69^ However, pasteurization showed no significant effect on recovery efficiency in our experiments (**Table S4**). Because paired samples processed by both concentrating methods underwent identical pasteurization treatment, this step does not affect the comparative conclusions of this study.

Incorporating immediate pasteurization after sample collection also allows workflows to transition rapidly to BSL-1 conditions, which provides a practical strategy for handling unknown or emerging pathogens such as during the early stages of the COVID-19 pandemic when transmission pathways were not yet fully understood. This approach may be particularly useful for laboratories with limited capacity to establish or maintain BSL-2 facilities, including those affiliated with wastewater treatment plants.

Ultrafiltration requires pretreatment steps such as centrifugation and filtration to remove solids and large aggregates, which may lead to the loss of viral RNA associated with sewage sludge.^29,70^ In a subset of samples, SARS-CoV-2 RNA was detected in pellets generated during the initial clarification step, indicating that part of the viral signal remained associated with solids typically discarded prior to ultrafiltration (**Table S6**). Similar observations have been reported in previous studies, where excluding solids from wastewater samples reduced viral detection rates.^71^ Detection of viral RNA in these pellets also confirms that TRIzol-chloroform extraction can recover viral RNA from solid fractions. Because freeze-drying concentrates both liquid and solid phases, viral RNA associated with both fractions can be extracted together, while insoluble solids are removed during the centrifugal phase-separation step of RNA extraction.

Freeze-drying also offers advantages for RNA preservation and workflow flexibility. After pasteurization, samples are immediately frozen at −80 °C, and the concentrating process proceeds under ultra-low temperatures, minimizing RNA degradation in environments where RNases are ubiquitous. This improved preservation may also contribute to the higher recovery efficiency and detection sensitivity observed for freeze-drying. In contrast, ultrafiltration typically requires immediate processing after pasteurization to avoid additional freeze–thaw cycles, limiting scheduling flexibility and increasing operational burden when laboratory resources are constrained. Freeze-drying allows samples to remain stored at ultra-low temperatures until processing, effectively decoupling sample collection from concentrating and providing greater flexibility for routine surveillance workflows.

### 3.10. Labor, Training, and Biosafety

From an operational perspective, ultrafiltration involves multiple centrifugation, balancing, and liquid-handling steps, requiring substantial operator training, particularly when handling unknown or emerging pathogens. These procedures also prolong operator exposure to wastewater and may increase the risk of pathogen transmission. For example, when the transmission route of SARS-CoV-2 was not yet fully understood, our original IBC-approved protocol classified wastewater processing as BSL-2+ conditions. Due to the potential for generating aerosols, ultrafiltration required rigorous PPEs (double-layer disposable nitrile gloves, face shields, closed-front gowns, and powered air-purifying respirators), along with extensive decontamination during multi-step centrifugation. These requirements led to labor-intensive workflows and limited throughput to approximately eight samples per day.

In contrast, although the freeze-drying method requires approximately three days to concentrate eight 200 mL samples to a final volume of ∼3 mL, it minimizes operator exposure. Sample handling is limited to brief steps during initial setup (e.g., cap loosening and flask mounting; **Figure 2**), after which the material remains sealed within a closed system equipped with multiple safeguards (**Figure 2**, **Section 2.2**). Decontamination is required only upon process completion. Moreover, freeze-drying involves substantially fewer liquid-handling steps, reducing opportunities for sample loss, cross-contamination, and other sources of human-factor variability throughout the concentrating process (**Section 2.2** and **2.3**).

## 4. Conclusion

In summary, using invaluable wastewater samples collected during the onset and waning phases of the first COVID-19 pandemic, this study indicates that freeze-drying may provide improved recovery and detection sensitivity compared with ultrafiltration under the conditions examined. The uniqueness and timing of these early-pandemic samples provide a rare opportunity to rigorously evaluate method performance under conditions of low and emerging viral prevalence. Freeze-drying’s higher recovery rates, detection ratios, and sensitivity for viral RNA are particularly crucial during such early phases, when viral loads are low and reliable detection is most challenging.

Freeze-drying’s operational advantages, including increased scheduling flexibility, reduced labor intensity, and minimized exposure risks, make it especially suitable for settings with limited technical expertise, personnel capacity, and biosafety infrastructure. Although freeze-drying requires a much longer processing time, the reduced need for continuous operator supervision and the use of inexpensive consumables can offset the slightly higher initial capital investment. This is particularly advantageous for newly established or developing laboratories with limited trained personnel or without extensive biosafety infrastructure to support frequent hands-on manipulation of pathogens.

This study represents a pivotal step in applying freeze-drying as a concentrating method for wastewater virus surveillance. Further optimization and comprehensive characterization are warranted, particularly in thoroughly evaluating the effects of pasteurization, mitigating matrix-associated inhibitors,^10^ developing approaches to normalize human contributions to wastewater,^72^ and systematic lead–lag time analysis.^73^ While this study focused on viral detection, future investigations should expand to other pathogens, including bacteria,^74,75^ fungi,^76,77^ and protozoa.^78,79^ Overall, our findings establish freeze-drying as a novel, sensitive, sustainable, and labor-saving alternative for wastewater concentrating, with strong potential for early warning and community-level surveillance of emerging pathogens.

## Data Availability

All data produced in the present work are contained in the manuscript

## ACKNOWLEDGMENT

During the preparation of this work, the author(s) utilized ChatGPT to only enhance readability and improve language. Following its use, the authors carefully reviewed and edited the content as necessary and took full responsibility for the final version of the publication. The authors would like to acknowledge Teresa T. Alcala and El Paso Water Laboratory personnel: Hugo Ruiz at John T. Hickerson Water Reclamation Facility, Martin L. Ortiz at Roberto Bustamante Wastewater Plant, Rick Dominguez at Haskell Street Wastewater Plant, Roberto Hernandez at Fred Hervey Water Reclamation, for their assistance during wastewater sample collection especially during the COVID-19 pandemic period. This work received funding from the College of Science, University of Texas at El Paso. The work has used instruments in the Border Biomedical Research Center (BBRC) that is supported by the National Institutes on Minority Health and Health Disparities (NIMHD) of the National Institutes of Health (NIH) under award number U54MD007592. Special thanks go to BBRC personnel Georgialina Rodriguez and Ana P. Betancourt who helped us use the instruments. RD and CX also received support from the National Institute of General Medical Sciences (NIGMS) of NIH under award number R01GM129525 and from the Welch Foundation under Grant Number AH-2126-20220331. WYL received support from NIGMS of NIH under the award number SC1CA245675. The content is solely the responsibility of the authors and does not necessarily represent the official views of the funding agencies.

