## SupplementalMaterials for "Freeze-Drying as a Novel Concentrating Method for Wastewater Detection of SARS-CoV-2"

### S1. Pre-filtration Safety Setup for Ultrafiltration Pre-clarification Step

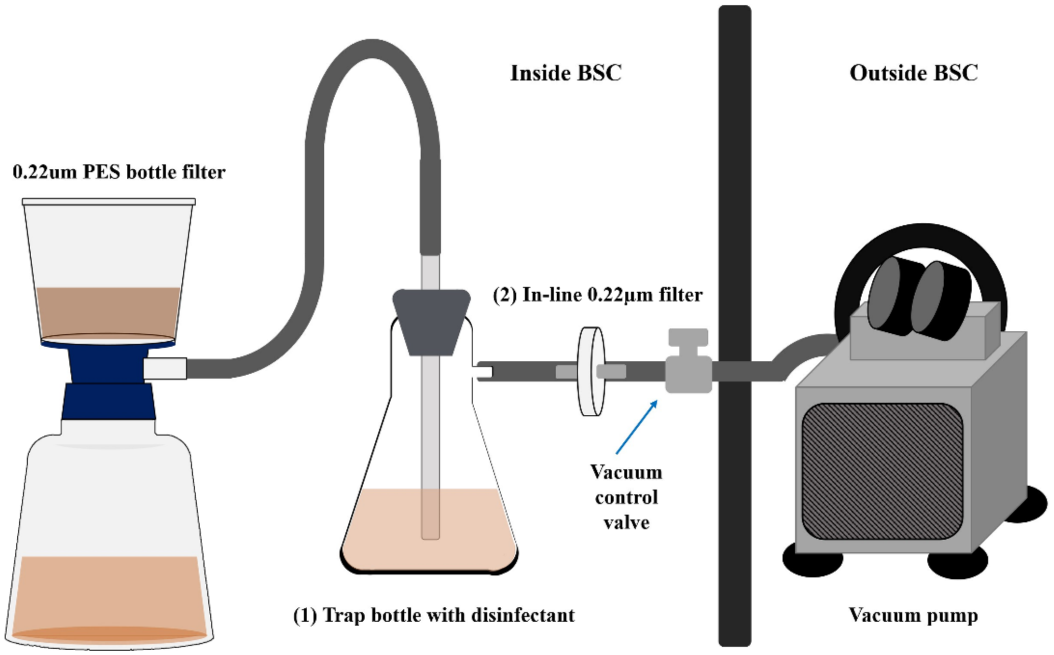


**Figure S1.** Schematic diagram of the in-line filter system. Key biosafety safeguard features are highlighted and labeled with numbers within the diagram.

### S2. Primer Information

**Table S1.** RT-qPCR primer sequences

| Assay | Primer | Sequence | Final concentration (nM) |
|---|---|---|---|
| | Forward primer | 5′-GACCCCAAAATCAGCGAAAT-3′ | 500 |
| SARS-CoV-2 N1 | Reverse primer | 5′-TCTGGTTACTGCCAGTTGAATCTG-3′ | 500 |
| | Probe | 5′-FAM-ACCCCGCATTACGTTTGGTGGACC-BHQ1-3′ | 125 |
| | Forward primer | 5′-TTACAAACATTGGCCGCAAA-3′ | 500 |
| SARS-CoV-2 N2 | Reverse primer | 5′-GCGCGACATTCCGAAGAA-3′ | 500 |
| | Probe | 5′-FAM-ACAATTTGCCCCCAGCGCTTCAG-BHQ1-3′ | 125 |
| | Forward primer | 5′-CGATGAGGCTATTCCGACTAGGT-3′ | 500 |
| OC43 | Reverse primer | 5′-CCTTCCTGAGCCTTCAATATAGTAACC-3′ | 750 |
| | Probe | 5′-6FAM-TCCGCCTGGCACGGTACTCCCT-BHQ-3′ | 50 |

### S3. RT-qPCR Primer-Probe Set Selection

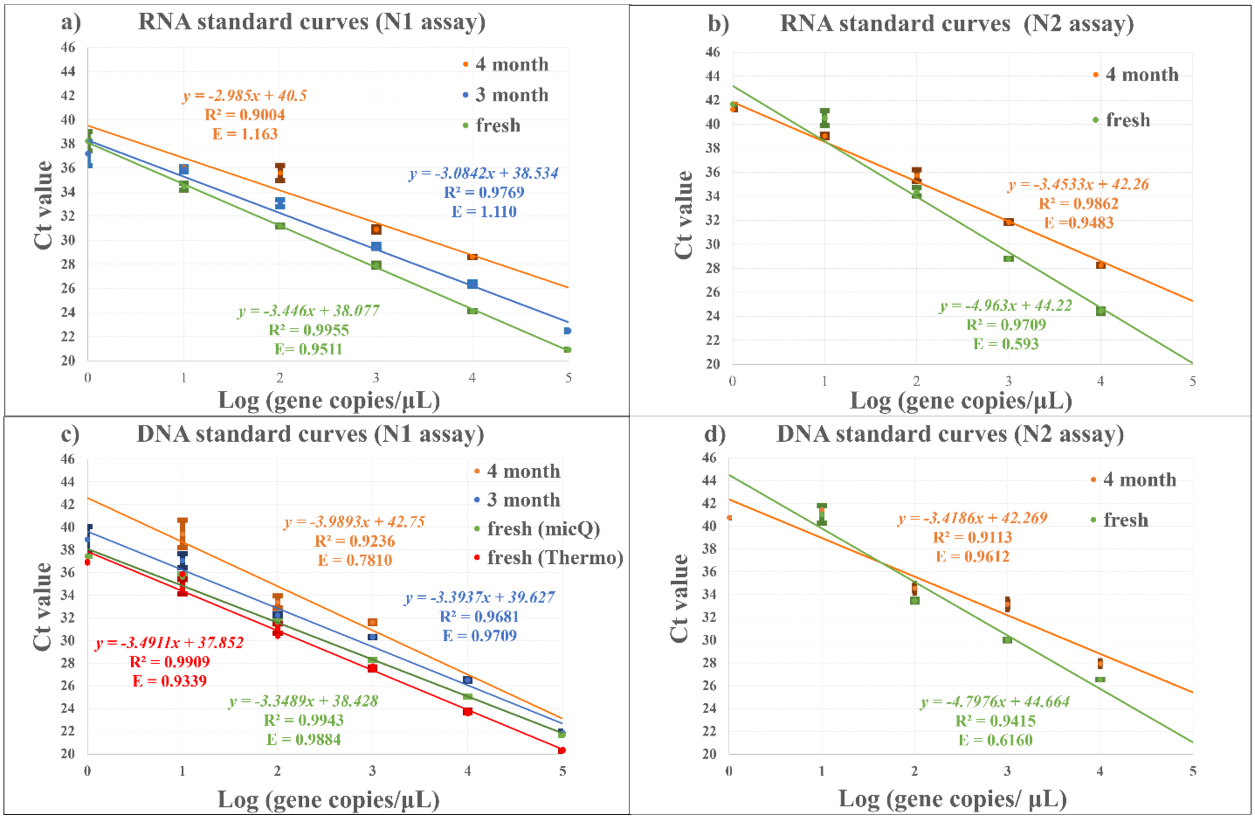


**Figure S2. RNA and DNA standard curves.** Panels a and b show fitted lines of RT-qPCR Ct mean values (y-axis), with error bars denoting ± SD, plotted against the logarithm of gene copy numbers (x-axis) of serially diluted RNA standards using the N1 and N2 primer–probe sets, respectively. Panels c and d present similar fitted lines for DNA standards. As indicated in the legend, different line colors represent standard curves generated from fresh standards or standards stored at −20 °C for 3 or 4 months. The fitted equations and corresponding $R^2$ values are shown in the same colors as their respective lines. Two fresh DNA standard curves are shown in Panel c: one generated using Bio Molecular Systems' Magnetic Induction Cycler (micPCR) and the other generated using the Thermo Fisher Scientific StepOnePlus™ Real-Time PCR System (Thermo).

**Table S2.** RNA and DNA standard curves information

| Primer-Probe Set (cycler) | | fresh | | | 3 month | | | 4 month | | |
|---|---|---|---|---|---|---|---|---|---|---|
| | | Equation (y=) | $R^2$ | Efficiency | Equation (y=) | $R^2$ | Efficiency | Equation (y=) | $R^2$ | Efficiency |
| **N1** | DNA(micQ) | -3.3489x + 38.428 | 0.9943 | 0.9884 | -3.3937x + 39.627 | 0.9681 | 0.9709 | -3.9893x + 42.75 | 0.9236 | 0.7810 |
| | RNA | -3.0842x + 38.534 | 0.9769 | 1.110 | -3.446x + 38.077 | 0.9955 | 0.9511 | -2.985x + 40.5 | 0.9004 | 1.163 |
| | DNA(Thermo) | -3.4911x + 37.852 | 0.9909 | 0.9884 | | | | | | |
| **N2** | DNA | -4.7976x + 44.664 | 0.9415 | 0.6160 | | | | -3.4186x + 42.269 | 0.9113 | 0.9612 |
| | RNA | -4.963x + 44.22 | 0.9709 | 0.593 | | | | -3.4533x + 42.26 | 0.9862 | 0.9483 |

Standard curve analysis in **Figure S2** and **Table S2** shows that across all points of RNA and DNA standards, the N1 primer-probe set consistently exhibited higher $R^2$ values, lower Ct values, and better efficiency than that of the N2 primer-probe set, indicating a stronger linear correlation between the measured and expected quantities. Additionally, the fitted lines of N1 for both RNA and DNA are lower than those of N2, reflecting overall lower Ct values for the N1 primer-probe set. These higher $R^2$ values and lower overall Ct values highlight the superior quantification accuracy and reliability of the N1 primer-probe set, making it the preferred choice.

However, the impact of standard degradation over time cannot be overlooked, as evidenced by the progressive increase in the heights (Ct values) of fitting lines observed for both RNA and DNA standards. Obviously, RNA exhibited more pronounced degradation than DNA, underscoring its higher susceptibility to degradation. The crossing observed in the RNA and DNA standard curves using N2 primer-probe set after storage in −20 °C for 4 months (**Figure S2b** and **Figure S2d**) suggests that the N2 primer-probe region may be more susceptible to degradation, resulting in deviations from the expected slope (approximately −3.3 to −3.4)[1] of the standard curve.

Given the superior linearity and stability of the DNA standard with the N1 assay, even after four months at −20 °C, it was used to estimate viral RNA copy numbers in wastewater concentrates for the main analyses.

While RNA standards are ideal for absolute RNA quantification, our study focuses on a relative comparison between two concentrating methods using identical calibration procedures.

Therefore, any systematic bias associated with DNA-based calibration would affect both methods equally and would not alter the comparative conclusions. Because of the higher degradation observed in RNA standards and the higher amplification efficiency of DNA standards, the DNA standard was selected for the comparative analyses in this study.

### S4. Standard Curve and RNA Quantification Calculation

The standard curve was plotted as RT-qPCR Ct value ($y$-axis) against the log of copy number ($x$-axis) estimated from the dilution of the original standards (**Figure S2**). Linear least-squares fitting was performed for each set of plotted data using the equation $y = a \times x + b$. The quality of these fittings was assessed using the coefficient of determination $R^2$ values. The slope $a$ should be negative, with an absolute value between 3.3 and 3.4, as the literature suggests that each 10-fold decrease in template concentration increases the Ct value by approximately three cycles.[2] Any value out of this range indicates potential issues with the dilution of standard templates or suboptimal RT-qPCR conditions. The $y$-intercept $b$ represents the Ct value when there is no template, where the fluorescence may arise from one or more of the following: non-specific amplification, primer-dimer formation, and background signal.[3]

According to the literature, the Ct value from RT-qPCR is linear to the logarithm of sample concentration.[1] Therefore, using the 10-fold diluted standard curves, Ct values from specific RT-qPCR can be converted to virus concentration in units of copies/mL using a fitted function of the DNA standard curve using N1 assay, as shown in **Figure S2**. Then, for each elution, the amount of virus detected can be calculated using the equation:

$$Virus\ detected\ (copies) = Virus\ concentration \times elution\ volume\ (30\mu L\ for\ 1^{st}\ elution\ and\ 100\mu L\ for\ 2^{nd}\ elution) \quad \textbf{(Equation S4.1)}$$

Total copies of the virus detected (column Total copies in **Table S6**) will be calculated as the sum of the virus detected from both first and second elutions. The detected viral load in the original unconcentrated sample (column D.c. in **Table S6**) can be determined using the equation:

$$Detected\ viral\ load\ (copies/mL) = Total\ copies\ /\ original\ sample\ volume \qquad \textbf{(Equation S4.2)}$$

### S5. Virus recovery rate calculation using OC43

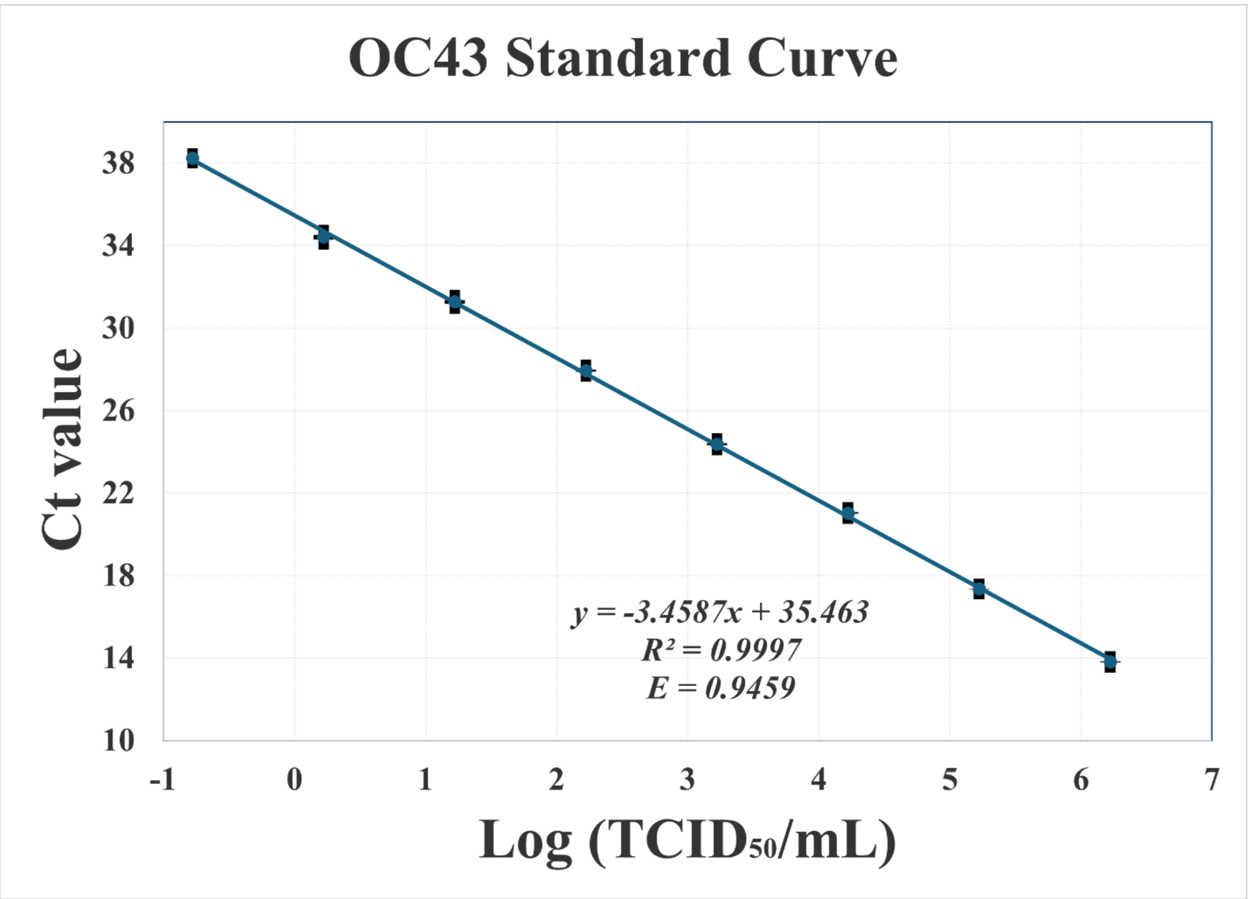


**Figure S3. OC43 standard curve.** qPCR mean Ct values (*y*-axis) with error bars denoting ± SD were plotted against the logarithm of virus concentration (*x*-axis) in units of $TCID_{50}$/mL, derived from serial dilutions of OC43 RNA standards. The fitted line illustrates the relationship using specific primers targeting OC43, with the corresponding equation and $R^2$ value shown on the graph.

For each experiment, a known quantity of OC43 virus (expressed in $TCID_{50}$ units) was spiked into 200 mL of wastewater. After concentration and RNA extraction, the recovered viral amount was quantified and expressed as the total $TCID_{50}$ equivalent recovered from the original 200 mL sample. Recovery efficiency (%) was calculated as:

$$Recovery\ Efficiency\ (\%) = \frac{Recovered\ Virus\ (TCID_{50})}{Initial\ Spiked\ Virus\ (TCID_{50})} \times 100 \quad \textbf{(Equation S5.1)}$$

**Table S3. Virus recovery efficiencies of freeze-drying and ultrafiltration with different procedures**

| **Spiking Level** | | | **Freeze Drying** | | **Ultrafiltration** | | | | |
|---|---|---|---|---|---|---|---|---|---|
| | Spiking time | Sample | Ct | Recovery efficiencies | Spiking time | Filter type | Sample | Ct | Recovery efficiencies |
| **High initial virus spiking** | Before pasteurization | F1 | 23.7±0.18 | 0.307±0.036% | Before pasteurization | 70mL 10k | U1 | 25.1±0.03 | 0.122±0.003% |
| | | F2 | 23.3±0.05 | 0.406±0.014% | | | U2 | 24.3±0.12 | 0.208±0.016% |
| | | F3 | 23.7±0.37 | 0.300±0.068% | | | U3 | 25.1±0.27 | 0.117±0.021% |
| | Average ± Std | | 23.6±0.31 | 0.338±0.065% | Average ± Std | | | 24.8±0.44 | 0.149±0.046% |
| **Low initial virus spiking** | After pasteurization | F4 | 27.7±0.28 | 0.108±0.020% | After pasteurization | 70mL 10k | U4 | 28.3±0.24 | 0.069±0.010% |
| | Before pasteurization | F5 | 27.3±0.05 | 0.135±0.005% | Before pasteurization | 70mL 10k | U5 | 28.8±0.06 | 0.051±0.002% |
| | | F6 | 27.6±0.49 | 0.120±0.037% | | 70mL 100k | U6 | 28.3±0.11 | 0.072±0.005% |
| | | F7 | 27.5±0.42 | 0.126±0.035% | | 70mL 100k | U7 | 28.6±0.17 | 0.059±0.007% |
| | Average ± Std | | 27.5±0.32 | 0.123±0.025% | Average ± Std | | | 28.5±0.26 | 0.062±0.011% |
| | | | | | | 15mL x 4 10K | U8 | 29.4±0.02 | 0.033±0.0005% |
| **No virus spiking** | - | F0 | Not detected | - | - | 70mL 10k | U0 | Not detected | - |
| **1 mL from initial virus spiking** | | F-1mL | Not detected | - | - | - | U-1mL | Not detected | - |

Virus recovery efficiencies obtained using ultrafiltration and freeze-drying were compared across different experimental parameters. A Wilcoxon rank-sum test was used to compare median recovery efficiencies between dosing levels and spiking-time conditions. The effect of filter type was subsequently evaluated within the low-dose ultrafiltration group. Because this comparison involved more than two filter types, a one-way analysis of variance (ANOVA) was used as an exploratory comparison of recovery efficiencies among groups. When significant differences were observed, a post hoc Tukey's Honest Significant Difference (HSD) test[4] was performed to identify pairwise differences between filters.

All statistical analyses were conducted in R (stats package, version 4.3.1)[5] using wilcox.test() for the Wilcoxon rank-sum test, aov() for ANOVA, and TukeyHSD() for post hoc comparisons. Results are summarized in **Table S4**.

**Table S4. Virus recovery efficiencies comparisons with different procedures.**

| Comparison | Recovery efficiencies (median [IQR Q1-Q3]) | | | p-value | Result |
|---|---|---|---|---|---|
| **High dosing: Freeze-drying vs. ultrafiltration** | 0.334% [0.321-0.391] | vs. | 0.125% [0.119-0.193] | $5.7 \times 10^{-4}$ | Significant |
| **Low dosing: Freeze-drying vs. ultrafiltration (70 mL filter)** | 0.128% [0.105-0.136] | vs. | 0.060% [0.052-0.069] | $1.7 \times 10^{-5}$ | Significant |
| **Freeze-drying: Spiking low dose before vs. after pasteurization** | 0.132% [0.122-0.139] | vs. | 0.105% [0.098-0.136] | 0.27 | Not significant |
| **Ultrafiltration: Spiking low dose before vs. after pasteurization** | 0.052% [0.049-0.065] | vs. | 0.074% [0.066-0.069] | 0.11 | Not significant |
| **Low dosing: Ultrafiltration filter cut-off size 10k vs. 100k** | 0.074% [0.066-0.069] | vs. | 0.067% [0.061-0.069] | 0.47 | Not significant |
| **Low dosing: Ultrafiltration 70 mL filter vs. 15 mL filter** | 0.060% [0.052-0.069] | vs. | 0.032% [0.032-0.033] | 0.004 | Significant |
| **Freeze-drying: High dosing vs. low dosing** | 0.334% [0.321-0.391] | vs | 0.128% [0.105-0.136] | $4.0 \times 10^{-6}$ | Significant |
| **Ultrafiltration: High dosing vs. low dosing (70 mL filter)** | 0.125% [0.119-0.193] | vs | 0.060% [0.052-0.069] | $4.0 \times 10^{-6}$ | Significant |

### S6. Comparison of Early Detection of Two Concentrating Methods

A Fisher's exact test was implemented to evaluate whether the proportion of positive detections differed between samples processed by freeze-drying and ultrafiltration. This test was chosen because the data consisted of categorical counts with small group sizes. The higher detection ratio ($p = 1.01 \times 10^{-5}$ using only the first elution; $p = 1.36 \times 10^{-6}$ when second elutions were also included) observed with the freeze-drying method indicates improved viral concentration performance compared with ultrafiltration under the conditions tested in this study. This analysis was performed using the fisher.test() function in R (stats package, version 4.3.1).[5]

### S7. Method-Specific Detection Limits

To further verify the method-specific detection limit defined above, wastewater samples were spiked with precise quantities of a SARS-CoV-2 DNA plasmid (IDT, Cat# 10006625) containing the complete nucleocapsid gene. Plasmid DNA with known copy numbers was added to wastewater samples and processed using the freeze-drying workflow to evaluate method-specific detection. The spiked samples were processed following the procedures described in **Sections 2.2** and **2.5**, and the results are summarized in **Table S5**.

To contextualize this sensitivity, the recovery efficiencies observed in this study (0.4%–0.1%; F2 and F4 in Table S3) imply that detecting 12 copies in the original sample would require approximately 3,000–12,000 copies to be present prior to concentration. This range reflects the inverse relationship between recovery efficiency and the initial viral quantity needed for detection. Notably, this inferred requirement is higher than the experimentally evaluated detection range using DNA plasmid (40–80 copies), although these estimates are based on different assumptions and should not be interpreted as directly equivalent measures of method sensitivity.

These experiments do not represent a full MIQE-compliant limit-of-detection (LOD) determination; rather, they provide partial verification of the magnitude of the method-specific detection limits defined in this study.

**Table S5.** Spiking at the limit

| Method | Spiking amount | Sample volume | Ct Values | | | |
|---|---|---|---|---|---|---|
| Freeze-drying | 40 copies | 200 mL | ND* | ND* | ND* | - |
| Freeze-drying | 80 copies | 200 mL | 35.4 | 35.6 | 35.1 | 34.7 |

* ND: Not Detected

### S8. Creatinine Measurement

Due to the substantial variation in wastewater flow rate caused by weather conditions (rainfall, drought, and temperature-dependent evaporation) and fluctuations in water usage throughout the day, estimating the proportion of human contribution to wastewater is challenging.[6] To address this issue, the concentration of creatinine, a human metabolic product, has been proposed as an internal control for assessing human contribution to wastewater.[7,8] Following published literature,[9] creatinine concentration was measured to evaluate human contribution to wastewater. Frozen pasteurized samples were thawed at 4°C, and 100 μL was aliquoted for creatinine analyses using a creatinine colorimetric detection assay (Enzo Life Sciences, Cat# ADI-907-030).

In this study, the creatinine levels detected were listed in the last column in **Table S6**. However, the measured creatinine concentration ranged from −6.489 mg/dL to 0.197 mg/dL, which is much lower than the sensitivity range (0.31 – 20 mg/dL). A review of the literature showed that reported creatinine levels in wastewater can be as low as 0.01 mg/dL,[10] are close to or outside the limit of the detection kit (0.042 mg/dL), raising concerns about the reliability of using creatinine as the internal standard for human contribution.

This discrepancy may be attributed to the dilution effect of non-human wastewater components, such as industrial and agricultural effluents, which can significantly impact creatinine concentrations, consistent with the literature.[11] When using pure water as blank, as recommended by the manufacturer, some of our measurements yielded negative values, suggesting the presence of certain interfering substances in wastewater that require further investigation.[12] As a result, creatinine concentrations were not used for normalization in this study because most measured values fell below the analytical sensitivity range of the assay, preventing reliable quantification. Other more accurate normalization assays will need to be investigated and developed in future studies.[13–18]

### S9. Virus detection data

**Table S6. Sample sites, dates, measurements, and calculations.**

| Site | Date | Freeze-drying | | | | | | | Ultrafiltration | | | | | | | | Creatinine conc. (mg/dL) |
|---|---|---|---|---|---|---|---|---|---|---|---|---|---|---|---|---|---|
| | | Sample volume (mL) | Ct Avg.* | | RNA copies* | | Total RNA copies | Detected Viral Load (cp/mL) | Sample volume (mL) | Ct Avg.** | | | RNA copies** | | Total copies*** | Detected Viral Load (cp/mL) *** | |
| | | | 1st | 2nd | 1st | 2nd | | | | 1st | 2nd | pt | 1st | 2nd | | | |
| | 4/14/20 | 140.8 | 33.1 | (36.5) | 692 | (240) | 931 | 6.62 | 140.8 | 36.2 | (36.3) | ND | 89 | (280) | 370 | 2.62 | - |
| | 4/16/20 | 192.6 | 34.6 | (35.4) | 251 | (510) | 761 | 3.95 | 190.2 | ND | ND | [41.0] | 0 | (0) | 0 | 0.00 | - |
| | 4/21/20 | 193.6 | 34.3 | (34.6) | 310 | (830) | 1139 | 5.89 | 192.5 | 31.0 | ND | ND | 2824 | (0) | 2824 | 14.67 | - |
| | 4/22/20 | 198.8 | 33.6 | (35.1) | 480 | (620) | 1100 | 5.53 | 196.0 | ND | ND | ND | 0 | (0) | 0 | 0.00 | -7.4 |
| | 4/23/20 | 193.7 | 42.0 | (35.6) | 2 | (453) | 455 | 2.35 | 199.3 | ND | ND | [36.3] | 0 | (0) | 0 | 0.00 | -5.0 |
| | 4/24/20 | 199.1 | 33.8 | (35.3) | 449 | (540) | 988 | 4.96 | 190.5 | ND | ND | ND | 0 | (0) | 0 | 0.00 | - |
| | 4/28/20 | 187.2 | - | - | - | - | - | - | 193.3 | - | - | - | - | - | - | - | - |
| | 4/29/20 | 189.9 | 31.4 | (33.3) | 2111 | (2061) | 4171 | 21.97 | 192.3 | ND | ND | [34.4] | 0 | (0) | 0 | 0.00 | -2.7 |
| | 4/20/20 | 193.7 | 38.1 | (36.9) | 26 | (182) | 208 | 1.07 | 188.2 | ND | ND | [33.4] | 0 | (0) | 0 | 0.00 | -0.5 |
| | 5/1/20 | 205.1 | 33.7 | ND | 463 | (0) | 463 | 2.26 | 189.5 | 34.8 | (36.4) | [35.5] | 230 | (265) | 495 | 2.61 | 0.1 |
| | 5/5/20 | 198.6 | 32.2 | (33.6) | 1245 | (1605) | 2851 | 14.35 | 188.1 | 33.4 | ND | [35.3] | 559 | (0) | 559 | 2.97 | -4.0 |
| HS | 5/6/20 | 206.5 | 31.8 | (33.8) | 1585 | (1427) | 3012 | 14.59 | 186.8 | ND | ND | ND | 0 | (0) | 0 | 0.00 | -4.1 |
| | 5/7/20 | 195.3 | 33.8 | (35.0) | 424 | (665) | 1089 | 5.58 | 180.3 | ND | ND | ND | 0 | (0) | 0 | 0.00 | -3.3 |
| | 5/8/20 | 191.2 | 34.6 | (36.9) | 262 | (182) | 444 | 2.32 | 174.4 | ND | ND | ND | 0 | (0) | 0 | 0.00 | -4.6 |
| | 5/12/20 | 204.3 | ND | ND | 0 | (0) | 0 | 0.00 | 189.5 | 36.5 | ND | [36.6] | 74 | (0) | 74 | 0.39 | -0.5 |
| | 5/13/20 | 214.2 | 38.2 | (36.9) | 24 | (185) | 209 | 0.98 | 161.6 | ND | ND | [36.1] | 0 | (0) | 0 | 0.00 | -1.3 |
| | 5/15/20 | 203.0 | ND | (31.1) | 0 | (8357) | 8357 | 41.17 | 183.1 | ND | ND | ND | 0 | (0) | 0 | 0.00 | - |
| | 5/27/20 | 195.7 | - | - | - | - | - | - | 188.9 | - | - | - | - | - | - | - | - |
| | 6/3/20 | 202.1 | ND | (32.4) | 0 | (3713) | 3713 | 18.37 | 184.5 | ND | (33.3) | ND | 0 | (2075) | 2075 | 11.25 | - |
| | 6/17/20 | 191.4 | 30.7 | (33.2) | 3440 | (2148) | 5588 | 29.20 | 190.7 | ND | ND | ND | 0 | (0) | 0 | 0.00 | -5.0 |
| | 7/1/20 | 190.7 | 30.4 | (34.1) | 3990 | (1204) | 5194 | 27.24 | 192.5 | ND | ND | ND | 0 | (0) | 0 | 0.00 | - |
| | 7/8/20 | 202.7 | 31.6 | (30.8) | 1902 | (10323) | 12226 | 60.31 | 186.7 | 37.5 | (37.0) | [36.3] | 38 | (181) | 219 | 1.17 | -3.4 |
| | 7/22/20 | 188.0 | 37.2 | ND | 46 | (0) | 46 | 0.25 | 185.9 | 35.8 | (32.4) | - | 113 | (3669) | 3782 | 20.34 | -1.4 |

**Table S6. Sample sites, dates, measurements, and calculations (continued)**

| Site | Date | Freeze-drying | | | | | | | Ultrafiltration | | | | | | | | Creatinine conc. (mg/dL) |
|---|---|---|---|---|---|---|---|---|---|---|---|---|---|---|---|---|---|
| | | Sample volume (mL) | Ct Avg.* | | RNA copies* | | Total RNA copies | Detected Viral Load (cp/mL) | Sample volume (mL) | Ct Avg.** | | | RNA copies** | | Total copies *** | Detected Viral Load (cp/mL) *** | |
| | | | 1st | 2nd | 1st | 2nd | | | | 1st | 2nd | pt | 1st | 2nd | | | |
| | 7/29/20 | 198.1 | 28.2 | (30.1) | 16989 | (16443) | 33432 | 168.76 | 181.9 | 30.9 | (34.9) | [29.4] | 2894 | (693) | 3587 | 19.72 | -3.6 |
| | 7/29/20 | 195.9 | - | (35.0) | - | (646) | 646 | 3.30 | 183.3 | 33.2 | (36.3) | - | 633 | (278) | 912 | 4.97 | -3.0 |
| | 8/5/20[a] | 212.0 | 30.4 | (31.8) | 4049 | (5475) | 9524 | 44.92 | 186.4 | 34.6 | (34.0) | [31.7] | 260 | (1247) | 1506 | 8.08 | -5.7 |
| | 8/5/20[b] | 209.4 | 30.5 | (31.3) | 3774 | (7600) | 11374 | 54.32 | 187.2 | 31.7 | (34.8) | [29.9] | 1779 | (742) | 2521 | 13.47 | -5.7 |
| | 8/10/20 | 253.5 | - | - | - | - | - | - | 195.1 | - | - | - | - | - | - | - | -3.1 |
| | 5/12/21[c] | 98.7 | 29.8 | (32.1) | 6155 | (4378) | 10533 | 106.77 | 91.9 | 37.5 | ND | - | 37 | (0) | 37 | 0.41 | -2.2 |
| HS | 5/12/21[d] | 98.7 | 29.7 | (32.5) | 6583 | (3443) | 10026 | 101.63 | 91.9 | 36.5 | (38.6) | - | 74 | (59) | 133 | 1.45 | - |
| | 5/12/21[e] | 100.2 | 29.3 | (32.5) | 8197 | (3446) | 11643 | 116.20 | 92.4 | 33.7 | (36.8) | - | 449 | (196) | 646 | 6.99 | - |
| | 5/12/21[f] | 100.2 | 29.3 | (32.1) | 8704 | (4476) | 13180 | 131.54 | 92.4 | 33.4 | (36.9) | - | 565 | (183) | 748 | 8.10 | - |
| | 5/20/21 | 192.5 | 36.4 | (34.3) | 78 | (1015) | 1093 | 5.68 | 191.0 | ND | (36.4) | [35.9] | 0 | (269) | 269 | 1.41 | - |
| | 5/20/21 | 189.6 | - | - | - | - | - | - | 193.4 | - | - | - | - | - | - | - | - |
| | 5/28/21 | 203.7 | ND | (34.8) | 0 | (742) | 742 | 3.64 | 236.4 | 32.8 | (35.5) | - | 860 | (479) | 1339 | 5.67 | -1.9 |
| | 5/28/21 | 204.8 | 37.8 | (32.2) | 30 | (4246) | 4277 | 20.88 | 281.1 | 32.5 | (34.7) | [31.5] | 1004 | (786) | 1789 | 6.37 | 0.3 |
| | 2/17/20 | 203.1 | 41.0 | (33.8) | 4 | (1479) | 1483 | 7.30 | 186.5 | 35.4 | ND | - | 151 | (0) | 151 | 0.81 | - |
| | 5/14/20 | 198.7 | 33.3 | (34.8) | 591 | (751) | 1342 | 6.76 | 189.0 | 35.2 | (35.7) | [35.4] | 176 | (405) | 581 | 3.08 | - |
| | 6/3/20 | 205.1 | 35.5 | (36.8) | 139 | (206) | 345 | 1.68 | 186.9 | 36.6 | ND | [36.5] | 67 | (0) | 67 | 0.36 | - |
| | 6/10/20 | 201.9 | 37.4 | (37.6) | 41 | (122) | 163 | 0.81 | 194.2 | - | ND | - | - | (0) | 0 | 0.00 | -2.4 |
| | 6/17/20 | 199.9 | 35.9 | (35.6) | 112 | (444) | 556 | 2.78 | 199.0 | ND | ND | [34.8] | 0 | (0) | 0 | 0.00 | -4.3 |
| JH | 6/24/20 | 198.1 | 35.3 | (35.0) | 162 | (675) | 837 | 4.22 | 199.5 | ND | (35.7) | [33.3] | 0 | (419) | 419 | 2.10 | -4.3 |
| | 7/3/20 | 104.5 | 31.4 | (32.3) | 2152 | (3769) | 5921 | 56.66 | - | - | - | - | - | - | - | - | - |
| | 7/8/20 | 210.9 | 35.2 | (35.2) | 177 | (576) | 753 | 3.57 | 189.0 | 36.4 | (30.5) | - | 77 | (13189) | 13266 | 70.19 | 0.2 |
| | 7/15/20 | 192.8 | 29.6 | (31.0) | 6825 | (9260) | 16085 | 83.43 | 199.9 | 35.2 | (35.6) | [31.2] | 167 | (456) | 623 | 3.12 | -5.2 |
| | 5/12/21 | 198.4 | 31.8 | (31.8) | 1664 | (5554) | 7218 | 36.38 | 187.5 | 32.8 | (35.8) | - | 844 | (393) | 1237 | 6.59 | - |
| | 5/20/21 | 201.8 | 39.0 | (34.1) | 15 | (1153) | 1167 | 5.79 | 289.1 | 29.6 | (32.5) | [33.6] | 7048 | (3500) | 10547 | 36.48 | - |

**Table S6. Sample sites, dates, measurements, and calculations (continued)**

| Site | Date | Freeze-drying | | | | | | | Ultrafiltration | | | | | | | | Creatinine conc. (mg/dL) |
|---|---|---|---|---|---|---|---|---|---|---|---|---|---|---|---|---|---|
| | | Sample volume (mL) | Ct Avg.* | | RNA copies* | | Total RNA copies | Detected Viral Load (cp/mL) | Sample volume (mL) | Ct Avg.** | | | RNA copies** | | Total copies *** | Detected Viral Load (cp/mL) *** | |
| | | | 1st | 2nd | 1st | 2nd | | | | 1st | 2nd | pt | 1st | 2nd | | | |
| FH | 4/26/20 | 183.2 | 34.9 | (34.8) | 205 | (768) | 973 | 5.31 | 190.9 | ND | ND | ND | 0 | (0) | 0 | 0.00 | - |
| | 4/28/20 | 187.1 | - | - | - | - | - | - | 193.3 | - | - | - | - | - | - | - | - |
| | 4/30/20 | 188.4 | 32.7 | (34.2) | 908 | (1115) | 2023 | 10.74 | 183.6 | 39.7 | (34.2) | [38.2] | 9 | (1086) | 1094 | 5.96 | -0.9 |
| | 5/5/20 | 212.2 | 39.2 | ND | 12 | (0) | 12 | 0.06 | 186.9 | 35.5 | ND | [36.8] | 144 | (0) | 144 | 0.77 | -1.9 |
| | 5/7/20 | 189.6 | 34.8 | (36.8) | 223 | (203) | 426 | 2.25 | 180.0 | 36.7 | ND | [35.3] | 65 | (0) | 65 | 0.36 | - |
| | 5/12/20 | 194.6 | - | - | - | - | - | - | 160.9 | - | - | - | - | - | - | - | -0.1 |
| | 5/14/20 | 199.0 | 32.3 | (34.9) | 1202 | (717) | 1919 | 9.64 | 183.1 | 35.9 | (36.2) | [33.1] | 110 | (295) | 404 | 2.21 | - |
| | 5/27/20 | 194.9 | 33.3 | (32.8) | 598 | (2767) | 3365 | 17.27 | 182.5 | 35.5 | ND | [33.2] | 142 | (0) | 142 | 0.78 | - |
| | 6/3/20 | 199.8 | 33.5 | (35.8) | 529 | (390) | 919 | 4.60 | 189.3 | ND | ND | [35.8] | 0 | (0) | 0 | 0.00 | - |
| | 6/10/20 | 175.7 | 33.1 | (36.1) | 670 | (307) | 977 | 5.56 | 196.0 | ND | ND | [35.1] | 0 | (0) | 0 | 0.00 | - |
| | 6/17/20 | 224.2 | ND | ND | 0 | (0) | 0 | 0.00 | 198.3 | ND | ND | ND | 0 | (0) | 0 | 0.00 | -3.4 |
| | 6/24/20 | 162.3 | ND | (31.4) | 0 | (7221) | 7221 | 44.49 | 198.0 | ND | ND | [30.2] | 0 | (0) | 0 | 0.00 | - |
| | 7/8/20 | 205.2 | 33.6 | (34.7) | 500 | (808) | 1308 | 6.37 | 188.1 | 34.4 | (39.0) | [37.1] | 285 | (47) | 332 | 1.77 | - |
| | 7/15/20 | 198.7 | 29.5 | (30.9) | 7312 | (10016) | 17328 | 87.21 | 186.4 | 35.5 | ND | [34.3] | 144 | (0) | 144 | 0.77 | - |
| | 7/22/20 | 184.7 | 33.1 | (32.5) | 709 | (3512) | 4221 | 22.85 | 200.8 | ND | (35.7) | [32.5] | 0 | (423) | 423 | 2.11 | -4.0 |
| | 5/12/21 | 202.7 | 31.2 | (31.9) | 2389 | (5214) | 7603 | 37.51 | 181.8 | 31.2 | (32.8) | - | 2461 | (2803) | 5264 | 28.95 | - |
| | 5/28/21 | 209.2 | 34.2 | (33.5) | 340 | (1771) | 2111 | 10.09 | 239.5 | 31.2 | (32.6) | - | 2370 | (3191) | 5561 | 23.22 | - |
| BU | 4/16/20 | 190.6 | 34.9 | (36.6) | 212 | (225) | 436 | 2.29 | 190.0 | ND | ND | [35.6] | 0 | (0) | 0 | 0.00 | - |
| | 4/22/20 | 198.7 | 32.1 | ND | 1353 | (0) | 1353 | 6.81 | 195.9 | 36.4 | (37.2) | [34.2] | 79 | (155) | 235 | 1.20 | -1.6 |
| | 4/24/20 | 191.0 | 34.4 | (35.9) | 293 | (371) | 664 | 3.48 | 190.9 | 33.5 | ND | [35.7] | 537 | (0) | 537 | 2.81 | - |
| | 4/29/20 | 191.5 | 30.4 | (34.1) | 4114 | (1157) | 5271 | 27.52 | 192.8 | ND | ND | [34.3] | 0 | (0) | 0 | 0.00 | -1.2 |
| | 5/1/20 | 211.3 | 35.2 | (33.1) | 173 | (2372) | 2546 | 12.05 | 187.7 | ND | ND | [30.0] | 0 | (0) | 0 | 0.00 | -0.4 |
| | 5/6/20 | 199.3 | 33.6 | (35.2) | 484 | (570) | 1054 | 5.29 | 187.0 | ND | ND | [34.6] | 0 | (0) | 0 | 0.00 | -2.8 |
| | 5/8/20 | 191.3 | 35.1 | (36.4) | 189 | (261) | 450 | 2.35 | 175.6 | ND | ND | [32.5] | 0 | (0) | 0 | 0.00 | -3.0 |

**Table S6. Sample sites, dates, measurements, and calculations (continued)**

| Site | Date | Freeze-drying | | | | | | | Ultrafiltration | | | | | | | | Creatinine conc. (mg/dL) |
|---|---|---|---|---|---|---|---|---|---|---|---|---|---|---|---|---|---|
| | | Sample volume (mL) | Ct Avg.* | | RNA copies* | | Total RNA copies | Detected Viral Load (cp/mL) | Sample volume (mL) | Ct Avg.** | | | RNA copies** | | Total copies *** | Detected Viral Load (cp/mL) *** | |
| | | | 1st | 2nd | 1st | 2nd | | | | 1st | 2nd | pt | 1st | 2nd | | | |
| BU | 5/13/20 | 199.7 | 31.3 | (31.1) | 2256 | (8800) | 11055 | 55.37 | 164.3 | 30.8 | ND | ND | 3138 | (0) | 3138 | 19.10 | - |
| | 5/15/20 | 202.0 | 36.4 | ND | 79 | (0) | 79 | 0.39 | 184.7 | ND | ND | [33.6] | 0 | (0) | 0 | 0.00 | - |
| | 5/20/20 | 191.2 | 35.6 | (33.7) | 133 | (1537) | 1670 | 8.73 | 193.7 | ND | ND | [31.1] | 0 | (0) | 0 | 0.00 | - |
| | 5/24/20 | 203.2 | - | - | - | - | - | - | 230.7 | - | - | - | - | - | - | - | - |
| | 6/3/20 | 198.0 | ND | (31.5) | 0 | (6562) | 6562 | 33.14 | 185.7 | ND | (35.1) | ND | 0 | (621) | 621 | 3.34 | - |
| | 6/17/20 | 191.6 | 30.1 | (33.0) | 4903 | (2515) | 7418 | 38.72 | 191.6 | 31.6 | (34.0) | [34.5] | 1841 | (1235) | 3076 | 16.05 | -2.8 |
| | 7/1/20 | 188.8 | 31.8 | (35.3) | 1623 | (528) | 2152 | 11.40 | 190.9 | 35.0 | (37.5) | [30.5] | 196 | (123) | 319 | 1.67 | - |
| | 7/8/20 | 190.8 | 33.4 | (34.1) | 558 | (1193) | 1751 | 9.18 | 191.8 | 34.6 | ND | ND | 260 | (0) | 260 | 1.35 | - |
| | 8/10/20 | 202.5 | 32.0 | (37.3) | 1434 | (147) | 1581 | 7.81 | 198.5 | 36.0 | ND | [34.4] | 102 | (0) | 102 | 0.52 | -3.2 |
| | 5/12/21 | 197.4 | 28.8 | (30.3) | 11495 | (14631) | 26126 | 132.35 | 188.0 | 30.6 | (33.2) | - | 3691 | (2156) | 5847 | 31.10 | - |
| | 5/28/21 | 206.7 | ND | (35.8) | 0 | (381) | 381 | 1.84 | 270.7 | 34.0 | (35.2) | - | 376 | (559) | 935 | 3.45 | - |
| Min | 2/17/20 | 98.7 | 28.2 | (30.1) | 0 | (0) | 0 | 0.00 | 91.9 | 29.6 | (30.5) | [29.4] | 0 | (0) | 0 | 0.00 | -7.4 |
| Max | 5/28/21 | 253.5 | 42.0 | (37.6) | 16989 | (16443) | 33432 | 168.76 | 289.1 | 39.7 | (39.0) | [41.0] | 7048 | (13189) | 13266 | 70.19 | 0.3 |
| Avg | | 191.6 | 33.5 | (34.0) | 1744 | (2496) | 4217 | 24.78 | 188.2 | 34.3 | (35.2) | [34.2] | 506 | (574) | 1073 | 5.44 | -2.8 |
| Std | | 26.5 | 3.0 | (2.0) | 3014 | (3441) | 5975 | 36.25 | 30.8 | 2.3 | (1.9) | [2.4] | 1141 | (1698) | 2269 | 10.99 | 1.9 |
| Median | | 198.1 | 33.5 | (34.1) | 472 | (830) | 1483 | 7.30 | 189.0 | 34.7 | (35.6) | [34.4] | 74 | (0) | 185 | 1.19 | -3.0 |
| IQR Q1 | | 190.9 | 31.4 | (32.4) | 87 | (376) | 703 | 3.52 | 184.7 | 32.8 | (34.0) | [32.8] | 0 | (0) | 0 | 0.00 | -4.1 |
| IQR Q3 | | 202.7 | 35.1 | (35.3) | 1843 | (3479) | 5755 | 31.17 | 193.4 | 36.0 | (36.4) | [35.7] | 285 | (416) | 871 | 5.49 | -1.4 |
| Non-zero minimum detection concentration (copies/mL): | | | | | | | | 0.06 | Non-zero minimum detection concentration (copies/mL) | | | | | | | 0.36 | - |

HS, Haskell R. Street Wastewater Treatment Plant; JH, John T. Hickerson Water Reclamation Facility; FH, Fred Hervey Water Reclamation Plant; BU, Roberto Bustamante Wastewater Treatment Plant; Data from the early stages of the COVID-19 pandemic (April 2020 to May 2020) were shaded in light grey.

a: sample was collected before ferric treatment.

b: sample was collected after ferric treatment.

c: sample was collected before ferric treatment and half of the sample was processed with the protocol described.

d: sample was collected before ferric treatment and half of the sample was processed with the protocol described but with more washes before eluting viral RNA.

e: sample was collected after ferric treatment and half of the sample was processed with the protocol described.

f: sample was collected after ferric treatment and half of the sample was processed with the protocol described but with more washes before eluting viral RNA.

*: numbers below are listed in two columns from 1st and 2nd elutions, respectively.

**: Numbers are listed in three columns from 1st and 2nd elutions as well as pellets (pt), respectively.

***: Numbers in columns do not include pellets because in early samples, pellets were not collected. In addition, our study is to compare ultrafiltration, not the pelleting method.

ND: not detected; - : data not collected (not detected will be treated as 0 in statistical test).

### S10. Correlation Between Wastewater Viral Load and COVID-19 Case Numbers

**Table S7. COVID-19 New case number and detected virus load via Freeze-drying and Ultrafiltration.**

| Date | New Cases | Detected virus load via Freeze-drying (copies/mL) | Detected virus load via Ultrafiltration (copies/mL) |
|---|---|---|---|
| 4/14/2020 | 10 | 6.62 | 2.62 |
| 4/16/2020 | 14 | 3.95 | 0.00 |
| 4/20/2020 | 28 | 1.07 | 0.00 |
| 4/21/2020 | 9 | 5.89 | 14.67 |
| 4/22/2020 | 3 | 5.53 | 0.00 |
| 4/23/2020 | 10 | 2.35 | 0.00 |
| 4/24/2020 | 8 | 4.96 | 0.00 |
| 4/29/2020 | 2 | 21.97 | 0.00 |
| 5/1/2020 | 7 | 2.26 | 2.61 |
| 5/5/2020 | 9 | 14.35 | 2.97 |
| 5/6/2020 | 1 | 14.59 | 0.00 |
| 5/7/2020 | 3 | 5.58 | 0.00 |
| 5/8/2020 | 10 | 2.32 | 0.00 |
| 5/12/2020 | 12 | 0.00 | 0.39 |
| 5/13/2020 | 7 | 0.98 | 0.00 |
| 5/15/2020 | 16 | 41.17 | 0.00 |
| 6/3/2020 | 11 | 18.37 | 11.25 |
| 6/17/2020 | 20 | 29.20 | 0.00 |
| 7/1/2020 | 27 | 27.24 | 0.00 |
| 7/8/2020 | 27 | 60.31 | 1.17 |
| 7/22/2020 | 172 | 0.25 | 20.34 |
| 7/29/2020 | 28 | 168.76 | 19.72 |
| 8/5/2020 | 108 | 44.92 | 8.08 |

(12) Xagoraraki, I.; O'Brien, E. Wastewater-Based Epidemiology for Early Detection of Viral Outbreaks. In *Women in Water Quality: Investigations by Prominent Female Engineers*;

O'Bannon, D. J., Ed.; Women in Engineering and Science; Springer International Publishing: Cham, 2020; pp 75–97. https://doi.org/10.1007/978-3-030-17819-2_5.
(13) Ribeiro, A. V. C.; Mannarino, C. F.; Novo, S. P. C.; Prado, T.; Lermontov, A.; de Paula, B. B.; Fumian, T. M.; Miagostovich, M. P. Assessment of crAssphage as a Biological Variable for SARS-CoV-2 Data Normalization in Wastewater Surveillance. *J. Appl. Microbiol.* **2024**, *135* (7), lxae177. https://doi.org/10.1093/jambio/lxae177.
(14) Cluzel, N.; Courbariaux, M.; Wang, S.; Moulin, L.; Wurtzer, S.; Bertrand, I.; Laurent, K.; Monfort, P.; Gantzer, C.; Guyader, S. L.; Boni, M.; Mouchel, J.-M.; Maréchal, V.; Nuel, G.; Maday, Y. A Nationwide Indicator to Smooth and Normalize Heterogeneous SARS-CoV-2 RNA Data in Wastewater. *Environ. Int.* **2022**, *158*, 106998. https://doi.org/10.1016/j.envint.2021.106998.
(15) Feng, S.; Roguet, A.; McClary-Gutierrez, J. S.; Newton, R. J.; Kloczko, N.; Meiman, J. G.; McLellan, S. L. Evaluation of Sampling, Analysis, and Normalization Methods for SARS-CoV-2 Concentrations in Wastewater to Assess COVID-19 Burdens in Wisconsin Communities. *ACS EST Water* **2021**, *1* (8), 1955–1965. https://doi.org/10.1021/acsestwater.1c00160.
(16) Foladori, P.; Cutrupi, F.; Cadonna, M.; Postinghel, M. Normalization of Viral Loads in Wastewater-Based Epidemiology Using Routine Parameters: One Year Monitoring of SARS-CoV-2 in Urban and Tourist Sewersheds. *J. Hazard. Mater.* **2024**, *478*, 135352. https://doi.org/10.1016/j.jhazmat.2024.135352.
(17) Hsu, S.-Y.; Bayati, M.; Li, C.; Hsieh, H.-Y.; Belenchia, A.; Klutts, J.; Zemmer, S. A.; Reynolds, M.; Semkiw, E.; Johnson, H.-Y.; Foley, T.; Wieberg, C. G.; Wenzel, J.; Johnson, M. C.; Lin, C.-H. Biomarkers Selection for Population Normalization in SARS-CoV-2 Wastewater-Based Epidemiology. *Water Res.* **2022**, *223*, 118985. https://doi.org/10.1016/j.watres.2022.118985.
(18) Isaksson, F.; Lundy, L.; Hedström, A.; Székely, A. J.; Mohamed, N. Evaluating the Use of Alternative Normalization Approaches on SARS-CoV-2 Concentrations in Wastewater: Experiences from Two Catchments in Northern Sweden. *Environments* **2022**, *9* (3), 39. https://doi.org/10.3390/environments9030039.
